# Single-cell analysis of bronchoalveolar cells in inflammatory and fibrotic post-COVID lung disease

**DOI:** 10.1101/2023.03.28.23287759

**Authors:** Puja Mehta, Blanca Sanz-Magallón Duque de Estrada, Emma K Denneny, Kane Foster, Carolin T Turner, Andreas Mayer, Martina Milighetti, Manuela Platé, Kaylee B Worlock, Masahiro Yoshida, Jeremy S Brown, Marko Z Nikolić, Benjamin M Chain, Mahdad Noursadeghi, Rachel C Chambers, Joanna C Porter, Gillian S Tomlinson

## Abstract

**Background:** Persistent radiological lung abnormalities are evident in many survivors of acute coronavirus disease 2019 (COVID-19). Consolidation and ground glass opacities are interpreted to indicate subacute inflammation whereas reticulation is thought to reflect fibrosis. We sought to identify differences at molecular and cellular level, in the local immunopathology of post-COVID inflammation and fibrosis.

**Methods:** We compared single-cell transcriptomic profiles and T cell receptor (TCR) repertoires of bronchoalveolar cells obtained from convalescent individuals with each radiological pattern, targeting lung segments affected by the predominant abnormality.

**Results:** Single-cell transcriptomes of inflammatory and fibrotic post-COVID lung disease closely resembled each other across all cell types. However, CD4 central memory T cells and CD8 effector memory T cells were significantly more abundant in those with inflammatory radiology. Clustering of similar TCRs from multiple donors was a striking feature of both phenotypes, consistent with tissue localised antigen-specific immune responses. There was no enrichment for known SARS-CoV-2-reactive TCRs, raising the possibility of T cell-mediated immunopathology driven by failure in immune self-tolerance.

**Conclusions:** We found no evidence that post-COVID radiographic inflammation and fibrosis are associated with differential immmunopathological pathways. Both show evidence of shared antigen-specific T cell responses, suggesting a role for therapies targeting T cells in limiting post-COVID lung damage.

## Introduction

Persistent functional and radiological lung abnormalities are evident at one year in approximately 20% of people who survive acute coronavirus disease 2019 (COVID-19) (Fabbri et al., 2023). Current understanding of the immunopathogenic mechanisms responsible for post-COVID lung disease (PCLD) is very limited (Mehta et al., 2022). Elevated numbers of airway CD4 and CD8 T cells have been reported (Cheon et al., 2021; Vijayakumar et al., 2022) and the post-COVID airway proteome displays evidence of ongoing epithelial injury that resolves with time (Vijayakumar et al., 2022). It is imperative to address this knowledge deficit to inform therapeutic interventions that could expedite resolution of pathology and minimize irreversible tissue damage, to reduce long term morbidity secondary to PCLD and the attendant burden on healthcare services.

There has been the impression of two major radiological patterns in PCLD: consolidation and ground glass opacities, thought to represent subacute inflammation, and reticulation, widely interpreted as fibrosis (Fabbri et al., 2023; Stewart et al., 2022). Inflammation predominates during acute COVID-19, but fibrosis is evident on 32% of computed tomography (CT) scans during hospitalisation. Follow-up imaging within the first year suggests radiological sequelae reduce with time, with less marked improvement in fibrosis (Fabbri et al., 2023). We hypothesised that these distinct radiological changes reflect distinct pathogenic mechanisms, which may require different treatments. We sought to evaluate the molecular characteristics of cellular function at the site of disease in PCLD, by single-cell RNA sequencing (scRNAseq) of bronchoalveolar cells from convalescent individuals infected during the first or second waves of the pandemic, with predominant CT features of inflammation or fibrosis at the time of sampling.

We showed that in comparison to fibrotic PCLD, the bronchoalveolar environment of inflammatory PCLD was enriched for CD4 T central memory cells (TCM) and CD8 T effector memory cells (TEM). Consistent with this finding, a higher proportion of CD4 TCM clones were expanded in the inflammatory phenotype. Both inflammatory and fibrotic PCLD bronchoalveolar T cells exhibited high levels of T cell receptor (TCR) clustering, indicative of an antigen-specific immune response, but there was no enrichment for known severe acute respiratory syndrome coronavirus 2 (SARS-CoV-2)-reactive sequences. No major differences were evident in any of the cell type-specific transcriptomic profiles of the two radiological phenotypes, suggesting that they may represent different manifestations of the same disease process.

## Results

### Increased abundance of bronchoalveolar T cells in inflammatory PCLD

Individuals undergoing bronchoscopy for clinical investigation of persistent respiratory symptoms and predominant radiological features of either inflammation or fibrosis following acute COVID-19, with no previous evidence of interstitial lung disease (ILD) were recruited (Figure 1a). Bronchoalveolar lavage (BAL) samples from lung segments affected by the predominant abnormality were obtained from five subjects with each radiological pattern (Figure 1a). Clinical and demographic characteristics are provided in Table 1 and Supplementary Table 1. Evaluation of the composition of post-COVID BAL by scRNAseq revealed that macrophages dominated in all subjects, with smaller T cell, NK T cell, dendritic cell, epithelial and B cell populations also identified (Figure 1b-d and Supplementary Figure 1a-d). Each of the cell types expressed high levels of independently established marker genes (Adams et al., 2020; Cheng et al., 2021; David et al., 2021; Davies et al., 2013; Davis and Wypych, 2021; Gardell and Parker, 2017; Glass et al., 2020; Murray and Wynn, 2011; Tsyklauri et al., 2023; van Aalderen et al., 2021; Villani et al., 2017), validating our annotations (Figure 1d). The notable difference between the two phenotypes was significantly higher abundance of CD4 T cells and CD8 T cells in inflammatory PCLD (Figure 1e). The relative proportions of the other seven cell types present were not found to differ between the two radiological phenotypes, providing confidence our analysis was not confounded by compositional bias, where alterations in the proportion of one cell type lead to many other cell types being falsely identified as differentially abundant.

**Figure 1.**
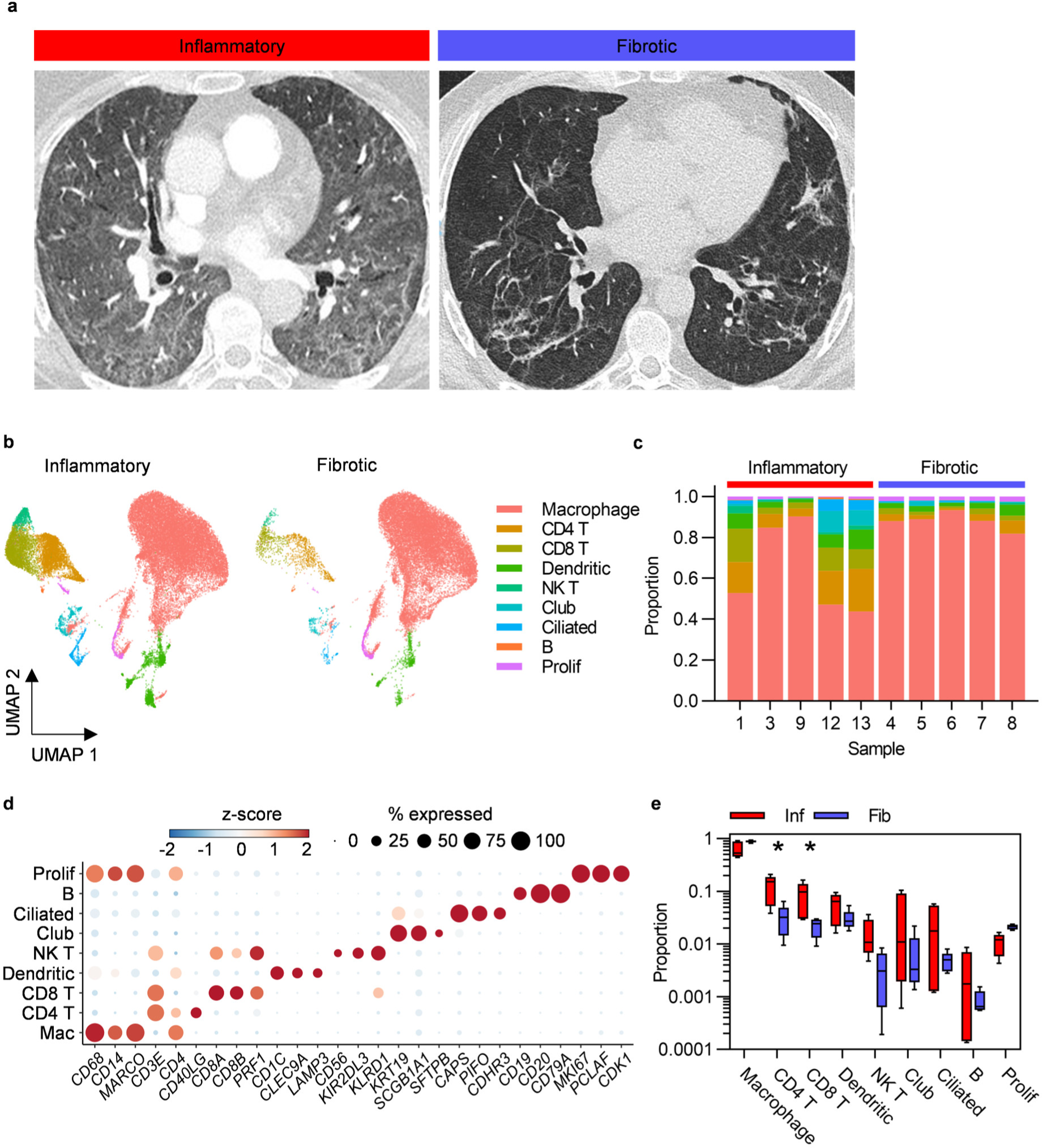
Higher abundance of bronchoalveolar T cells in inflammatory post-COVID-19 lung disease (PCLD). **a** Representative computed tomography (CT) images for each radiological phenotype. **b** Uniform manifold approximation and projection (UMAP) embedding of 55,776 bronchoalveolar single-cell transcriptomes obtained from five individuals with radiological features of pulmonary inflammation (33,553 cells) and five individuals with radiological features of pulmonary fibrosis (22,223 cells) following COVID-19, split by PCLD phenotype, color coded by cell type. Cell type annotation was achieved by assignment of Azimuth human lung reference gene signatures using the SCINA R package and in the case of dendritic cells and B cells, expression of literature-based markers. Prolif = proliferating cells, identified by their high module score for a gene signature representing the cellular proliferation response. **c** Cellular composition of each BAL sample defined by single-cell RNA sequencing (scRNAseq). Color indicates cell type and bar height represents proportion. **d** Dot plot visualization of the expression of independently established marker genes for each cell type; “Mac” = macrophage. Dot size represents the percentage of cells expressing the gene in each cell type, color shows the z-scores of average log-normalized expression for each cell type compared to the entire data set. **e** Comparison of the proportions of each cell type in inflammatory and fibrotic PCLD. Horizontal lines indicate median, box limits the interquartile range and whiskers the 5^th^ to 95^th^ percentiles, * = FDR<0.05.

**Table 1.**
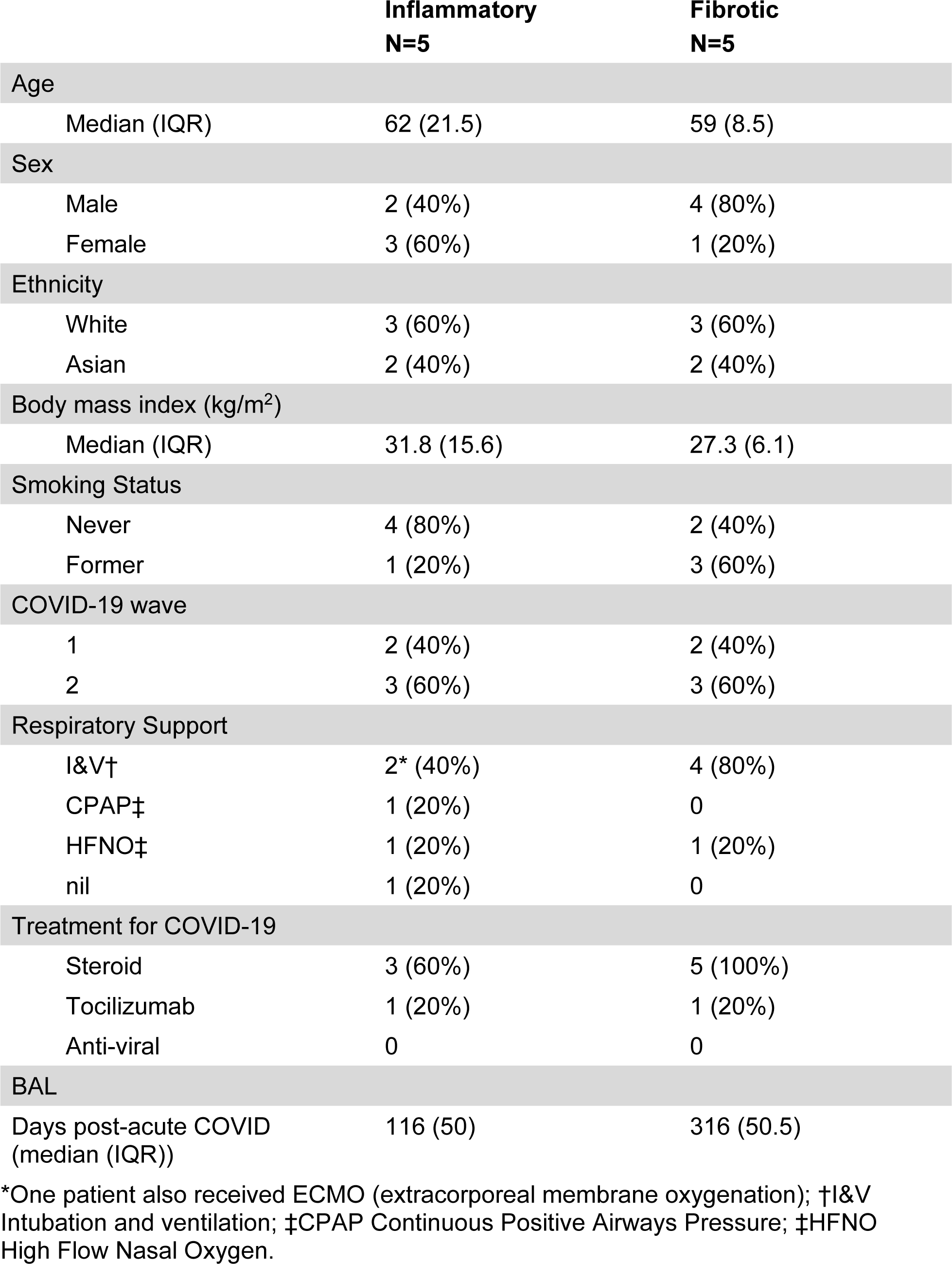
Clinical and demographic information summary.

|  | Inflammatory<br>N=5 | Fibrotic<br>N=5 |
| --- | --- | --- |
| Age |  |  |
| Median (IQR) | 62 (21.5) | 59 (8.5) |
| Sex |  |  |
| Male | 2 (40%) | 4 (80%) |
| Female | 3 (60%) | 1 (20%) |
| Ethnicity |  |  |
| White | 3 (60%) | 3 (60%) |
| Asian | 2 (40%) | 2 (40%) |
| Body mass index (kg/m<sup>2</sup>) |  |  |
| Median (IQR) | 31.8 (15.6) | 27.3 (6.1) |
| Smoking Status |  |  |
| Never | 4 (80%) | 2 (40%) |
| Former | 1 (20%) | 3 (60%) |
| COVID-19 wave |  |  |
| 1 | 2 (40%) | 2 (40%) |
| 2 | 3 (60%) | 3 (60%) |
| Respiratory Support |  |  |
| I&V† | 2* (40%) | 4 (80%) |
| CPAP‡ | 1 (20%) | 0 |
| HFNO‡ | 1 (20%) | 1 (20%) |
| nil | 1 (20%) | 0 |
| Treatment for COVID-19 |  |  |
| Steroid | 3 (60%) | 5 (100%) |
| Tocilizumab | 1 (20%) | 1 (20%) |
| Anti-viral | 0 | 0 |
| BAL |  |  |
| Days post-acute COVID<br>(median (IQR)) | 116 (50) | 316 (50.5) |

\*One patient also received ECMO (extracorporeal membrane oxygenation); †I&V Intubation and ventilation; ‡CPAP Continuous Positive Airways Pressure; ‡HFNO High Flow Nasal Oxygen.

To explore differences in cell type-specific transcriptional profiles between the two radiological phenotypes, we first aggregated gene expression count data for each cell type for each donor to form “pseudobulks”. We leveraged the ability of pseudobulk statistical approaches to account for variability of biological replicates, allowing detection of genuine differential gene expression whilst minimizing false discoveries (Squair et al., 2021). Very few cell type-specific differentially expressed genes were identified (Supplementary Table 2), which precluded further bioinformatic analysis and suggested the transcriptomes of inflammatory and fibrotic PCLD were similar for all cell types.

Detection of transcriptomic differences between the two groups using pseudobulk statistical analysis may have been limited by small sample size. Therefore, we repeated the comparison of inflammatory and fibrotic PCLD at the level of individual cells. Single-cell statistical analysis identified thousands of cell type-specific differentially expressed genes in each radiological phenotype. However, single-cell differential gene expression analysis has a propensity for false positive results (Squair et al., 2021). To mitigate against this, we sought to assess whether differentially expressed gene lists represented differentially enriched biological pathways in the two study groups. This was not evident at the level of biological processes or upstream regulator analysis (Supplementary Figure 2 and 3), which suggested enrichment of overlapping processes and pathways despite apparent differential gene expression. Hence, our single-cell analysis also supports the outcome of the pseudobulk analysis.

### CD4 central memory and CD8 effector memory are the predominant T cell subsets in PCLD

Given that greater abundance of T cells in inflammatory cases was the only robust difference between the two PCLD radiological phenotypes, we re-clustered these populations alone to undertake a more detailed analysis. This revealed CD4 TCM and CD8 TEM as the two predominant T cell subsets in the PCLD bronchoalveolar environment, and a smaller population of regulatory T cells (Treg). A small mixed T cell cluster, which expressed high levels of interferon-stimulated genes and an NK cell cluster were also present in all samples. A minor population of gamma delta T cells was identified in one individual with inflammatory PCLD (Figure 2a-c). There was no difference in the relative proportions of any T cell subset between the two PCLD phenotypes (Figure 2d).

**Figure 2.**
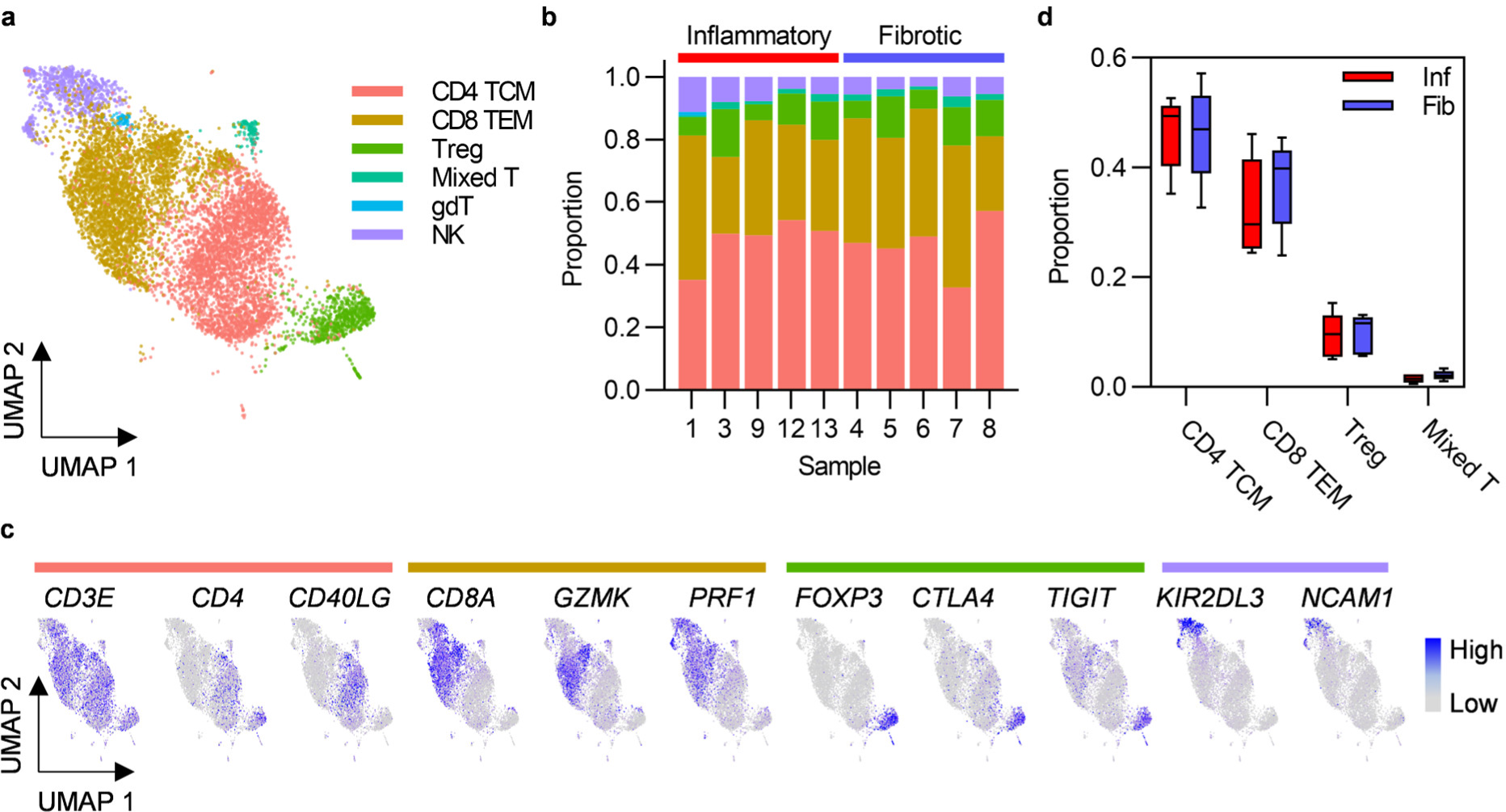
Post-COVID-19 bronchoalveolar T cells are dominated by CD4 central memory and CD8 effector memory subsets. **a** UMAP embedding of 9196 transcriptomes of T cells identified in Figure 1b, colored by cell type. T cell subset annotation was based on assignment of Azimuth human PBMC reference marker gene signatures and additional published CD4 T cell signatures by the SCINA R package. **b** Relative proportions of each T cell subset in each subject, colors represent cell type. **c** Feature plots demonstrating the expression of marker genes for the three principal T cell subsets and NK cells, colored by scaled, log-normalized counts, projected on to the T cell UMAP. **d** Comparison of the proportions of T cell subsets found in all individuals in inflammatory and fibrotic post-COVID-19 lung disease (PCLD). Horizontal lines indicate median, box limits the interquartile range and whiskers the 5^th^ to 95^th^ percentiles.

Analysis of T cell subset pseudobulks revealed no genes as differentially expressed in either group (Supplementary Table 2), consistent with our earlier observation of few differences between the transcriptional programmes of the two PCLD phenotypes at the level of broad cell types defined using the full dataset. In single-cell differential expression analysis of T cells, genes expressed at a significantly higher level in CD4 TCM and CD8 TEM in inflammatory PCLD exhibited weak enrichment for immune response signalling pathways (Supplementary Figure 4a). Robust enrichment of cellular pathways in anti-viral responses was evident for genes expressed at a significantly higher level in CD4 TCM and CD8 TEM in fibrotic PCLD (Supplementary Figure 4b), but this was not supported by enrichment for Type I interferon signalling in upstream regulator analysis (Supplementary Figure 5b). Interestingly, interleukin (IL)2 and IL15 were the most statistically significant upstream regulators of CD4 TCM and CD8 TEM differentially expressed genes in inflammatory PCLD but not fibrotic PCLD, consistent with the notion of cytokine driven proliferation leading to increased abundance of T cells in the former phenotype (Supplementary Figure 5a-c).

### Alveolar macrophage and monocyte subsets in PCLD are consistent with healthy airspace myeloid populations

Macrophages are the most abundant cell types in healthy airspaces (Mould et al., 2021) and were also found to be the most abundant cell type in PCLD BAL samples in the present study. Macrophage subpopulations have been implicated in the pathogenesis of fibrosis associated with severe COVID-19 and idiopathic pulmonary fibrosis (Nouno et al., 2019; Wendisch et al., 2021). We therefore re-clustered macrophages alone to further our understanding of macrophage subpopulations in PCLD. Two populations with similar expression of macrophage markers and comparable transcriptomic profiles, likely representative of the transcriptional spectrum of resident healthy alveolar macrophages (AM) (Mould et al., 2021) were combined for subsequent analysis (Figure 3a,b and Supplementary Figure 6a,b). We identified three further small specialized AM subsets previously detected in healthy individuals (Mould et al., 2021), characterized by high levels of proinflammatory molecule expression, “Inflam AM”, metal-binding metallothioneins, “MT-AM”, or interferon-stimulated genes “IFN stim AM”. Proliferating macrophages were delineated by high expression of a gene module representing the cellular proliferation response (Chandran et al., 2022) (Figure 3a-d).

**Figure 3.**
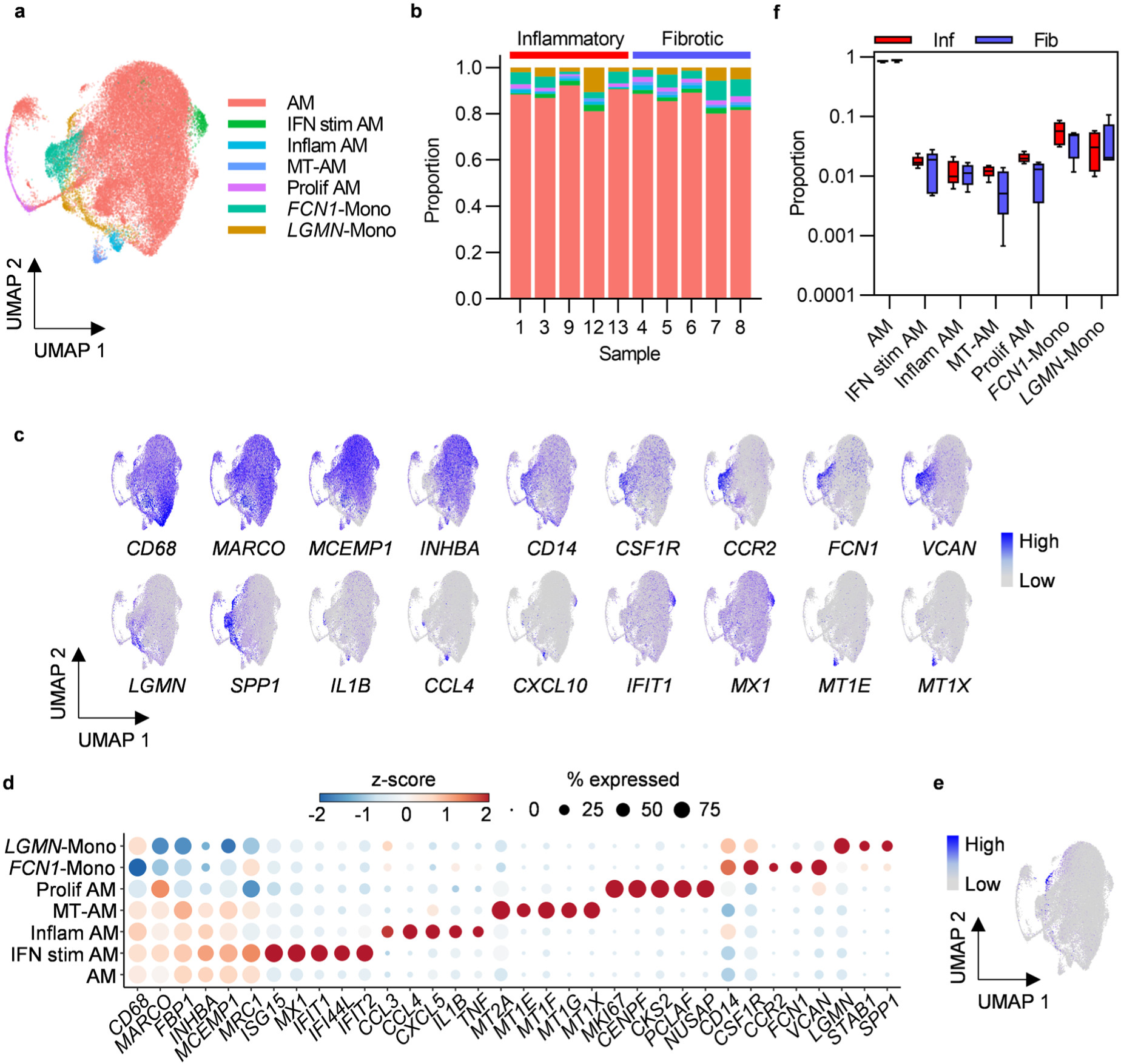
Bronchoalveolar macrophage and monocyte subsets in post-COVID-19 lung disease (PCLD). **a** UMAP embedding of 38,010 macrophage transcriptomes identified in Figure 1b, colored by cell type. Myeloid populations were annotated by assignment of Azimuth human lung reference marker genes and additional signatures derived from published literature using the SCINA R package and by visualising expression of canonical marker genes for some subsets. **b** Relative proportions of each myeloid subset within each individual, colors represent cell type. **c** Expression of marker genes for PCLD macrophage and monocyte populations, colored by scaled, log-normalized counts, projected on to the myeloid cell UMAP. **d** Dot plot visualization of the expression of selected marker genes for each macrophage and monocyte subset. Dot size represents the percentage of cells expressing the gene in each myeloid subset, color shows the z-scores of average log-normalized expression for each subset compared to the entire data set. **e** Expression of a profibrotic macrophage gene signature derived from idiopathic pulmonary fibrosis, calculated on a single-cell level, colored by module score and projected on to the macrophage UMAP. **f** Comparison of the proportions of bronchoalveolar myeloid populations in inflammatory and fibrotic PCLD. Horizontal lines indicate median, box limits the interquartile range and whiskers the 5^th^ to 95^th^ percentiles.

Monocyte-like cells are present in healthy airspaces, and suggested by trajectory inference and increasing expression of macrophage marker genes over pseudotime to differentiate into AM, implying constant trafficking of monocytes into the lung (Mould et al., 2021). Consistent with this, we identified *CD14, FCN1* expressing classical monocytes (*FCN1*-Mono) (Mould et al., 2021; Wauters et al., 2021) characterized by high levels of *CCR2*, the receptor for monocyte chemoattractant protein, suggestive of recent recruitment from the peripheral blood (Auffray et al., 2009) (Figure 3c,d). A second *LGMN* and *SPP1* expressing subset (*LGMN*-Mono), recently identified as a rare population defined by expression of cell-matrix interaction genes in healthy airspaces (Mould et al., 2021), was also present (Figure 3c,d). In contrast to acute severe COVID-19 where proinflammatory monocytes and profibrotic *SPP1, LGMN* expressing macrophages have been reported to be abundant (Liao et al., 2020; Wendisch et al., 2021), monocytes represented a minor constituent of PCLD BAL. Neither subset expressed high levels of inflammatory mediators (Figure 3c,d) and very few cells expressed a gene signature characteristic of profibrotic macrophages recently identified in idiopathic pulmonary fibrosis (Adams et al., 2020), (Figure 3e and Supplementary Figure 7a).

No differences in the relative proportions of any of the myeloid populations were evident between the two PCLD phenotypes (Figure 3f). Very few gene expression differences were detected between the two phenotypes by pseudobulk analysis of macrophage or monocyte subsets (Supplementary Table 2). Similar to the analysis of the full dataset stratified by broad cell type, re-clustering macrophages to identify more discrete sub-populations did not reveal transcriptomic differences between inflammatory and fibrotic PCLD.

Single-cell differential gene expression analysis revealed weak enrichment for immune response and cellular metabolism pathways in both groups (Supplementary Figure 7b,c). Minor enrichment for TGFβ-mediated signalling was evident for genes expressed at higher levels in AM, Inflam-AM and *FCN1*-Mono in inflammatory PCLD (Supplementary Figure 7b). Proinflammatory cytokines, T cell activation factors, SPP1 and TGFβ were identified as statistically enriched upstream regulators of differentially expressed genes in both monocyte populations in inflammatory PCLD (Supplementary Figure 8a,b,d,e). However, as for analysis at the level of broad cell types and refined T cell subsets, there was considerable overlap between molecules predicted to drive gene expression differences in each PCLD phenotype (Supplementary Figure 8c,f), suggesting that between group differential gene expression in this analysis did not represent differential biology between radiological patterns of disease.

### Highly related TCRs indicate antigen-specific immune responses in PCLD

To further evaluate T cell responses in PCLD we undertook single-cell TCR sequencing (scTCRseq) and compared the TCR repertoire in each radiological phenotype. We successfully obtained scTCRseq data for five fibrotic and three inflammatory cases. Increased T cell abundance in inflammatory PCLD may reflect increased numbers of unique T cell clones or increased expansion of individual clones. Expanded clonotypes identified by being present with a frequency of greater than one, were evident within the three major T cell subsets in both inflammatory and fibrotic PCLD (Figure 4a). A greater proportion of CD4 TCM clones were expanded in the inflammatory phenotype, consistent with our observation of the higher abundance of this subset in this group. However, the proportion of expanded CD8 TEM and Treg clones was similar in both phenotypes (Figure 4b).

**Figure 4.**
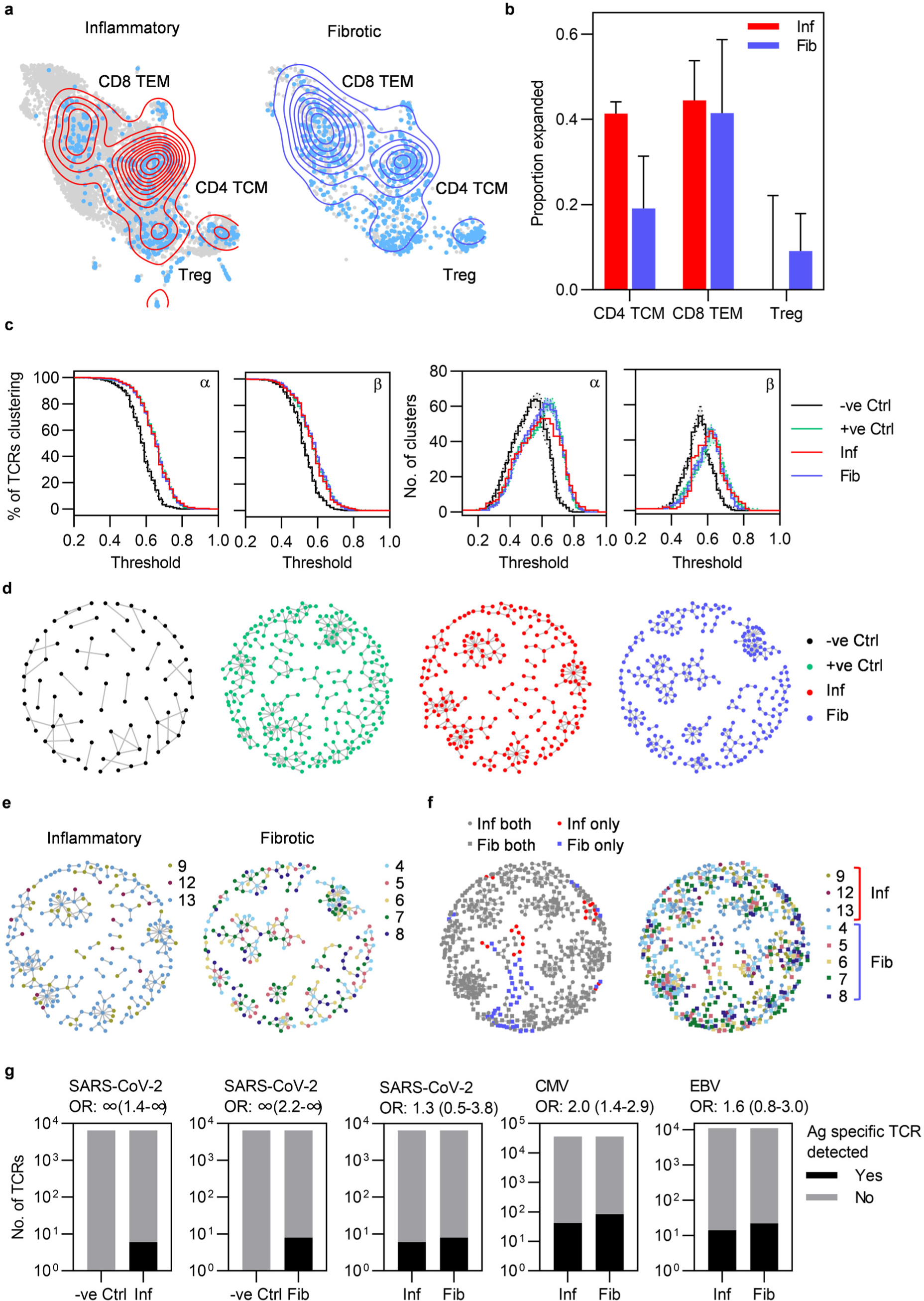
Highly similar T cell receptors (TCRs) characterize both inflammatory and fibrotic post-COVID lung disease (PCLD). **a** T cell clonal expansion is visualized on the T cell UMAP split by radiological PCLD phenotype. TCR sequences detected at a frequency of greater than one are colored light blue and contour lines provide a 2D representation of TCR density overlaid in red for inflammatory PCLD and blue for fibrotic PCLD. **b** Comparison of the proportion of expanded TCR sequences, defined as those detected at a frequency greater than one in the three largest T cell subsets identified in Figure 2a in the two PCLD phenotypes. **c** Percentage of complementarity determining region (CDR)3 alpha and beta chain amino acid sequences clustering and number of clusters generated over a range of thresholds above which two TCRs are considered similar, for inflammatory and fibrotic PCLD bronchoalveolar lavage (BAL) samples, negative control PBMC samples not expected to cluster highly and positive control PBMC samples known to cluster highly, analyzed separately. **d** Representative network diagrams of TCR β chain clusters present in each group described in (**c**). Nodes represent TCRs, related TCRs are connected by an edge and colors represent the groups. **e** Network diagrams of related TCR β chains in each PCLD phenotype in which nodes are colored by donor. **f** Network diagrams visualizing TCR β chain clusters identified by combined analysis of the two PCLD phenotypes. Nodes are colored by donor (right) or radiological phenotype and membership of clusters composed of one or both PCLD groups (left), circular nodes represent inflammatory PCLD and square nodes fibrotic PCLD. **g** Number of TCR sequences (α and β genes combined) annotated for SARS-CoV-2, cytomegalovirus (CMV), and Epstein-Barr virus (EBV) in VDJdb either detected or not detected in TCR sequences from individuals with each PCLD phenotype and negative controls, giving the odds ratio ±95% confidence interval (Fisher’s exact test) for enrichment of antigen-specific TCR sequences in each instance.

As T cell clonal expansion was evident in both PCLD phenotypes, we next sought to identify related TCRs for each group based on the similarity of their antigen specificity-determining complementarity determining region (CDR)3 amino acid sequences, on the premise that clusters of related TCRs recognize similar epitopes (details of the analysis are provided in the methods). We hypothesized that the inflammatory cases would exhibit more clustering than the fibrotic cases, given the trend towards a greater proportion of expanded CD4 TCM clones and increased abundance of both CD4 TCM and CD8 TEM in the inflammatory group. However, high levels of clustering were evident in both inflammatory and fibrotic PCLD. In benchmarking, this level of clustering was similar to that observed for expanded peripheral blood TCR clones detected following non-severe SARS-CoV-2 infection, and exceeded that in non-expanded TCRs from non-infected individuals from the same cohort (Chandran et al., 2022; Milighetti et al., 2023) (Figure 4c,d). In both radiological groups of PCLD the vast majority of clusters contained TCRs from multiple donors (Figure 4e) but there was minimal sharing of identical CDR3 sequences between different individuals (Supplementary Table 3). As a further comparison of inflammatory and fibrotic PCLD T cell repertoires we clustered CDR3 amino acid sequences from both groups together. Strikingly, most clusters contained TCRs from both inflammatory and fibrotic samples and multiple donors, suggesting the presence of T cell clones that recognize similar antigens across both phenotypes. A few small clusters composed uniquely of either inflammatory or fibrotic PCLD TCRs were evident, indicative of subtle differences between the two repertoires (Figure 4f).

The high level of relatedness between TCRs in PCLD is suggestive of antigen-specific immune responses. We therefore sought to determine whether these repertoires were enriched for T cells specific for SARS-CoV-2 by comparison to other common viruses. Of the SARS-CoV-2-reactive TCRs identified in the VDJdb database, six were present in TCR data from inflammatory PCLD and eight in fibrotic PCLD. None were identified in equivalent sized healthy peripheral blood repertoires. However, fewer SARS-CoV-2-specific-TCRs were detected than Epstein-Barr virus (EBV)-specific or cytomegalovirus (CMV)-specific sequences, indicating no enrichment for SARS-CoV-2-specific T cells at the site of disease. There was no enrichment for SARS-CoV-2-reactive or EBV-reactive TCRs in inflammatory compared to fibrotic PCLD. However, CMV-specific TCRs were significantly enriched in the fibrotic group (Figure 4g). TCRs found in clusters composed uniquely of either PCLD phenotype were not enriched for known virus reactive-sequences (Supplementary Table 4).

## Discussion

We report the first comparative molecular analysis of cells sampled by bronchoalveolar lavage from patients displaying either predominantly inflammatory or fibrotic pulmonary radiological sequelae following COVID-19. The bronchoalveolar environment of inflammatory PCLD was characterised by significantly increased abundance of CD4 central memory and CD8 effector memory T cells compared to fibrotic PCLD. Clustering of similar TCRs from different donors, far exceeding that observed in healthy peripheral blood, was evident in both radiological phenotypes, suggestive of an antigen-specific immune response localised to the lung. The transcriptomes of post-COVID radiological inflammation and fibrosis were highly similar for all bronchoalveolar cell types, dominated by cellular processes involved in inflammatory and immune responses. We found no robust evidence of enhanced activity of tissue damage or wound repair pathways in those with fibrotic radiological changes. Although many cell type-specific differentially expressed genes were identified between the two radiological phenotypes by single-cell methods, there were no systematic differences in enriched biological pathways or their upstream regulators among these differentially expressed genes. Furthermore, almost no differences in gene expression between the fibrotic and the inflammatory groups were detected if gene expression within cell types was aggregated by donor before differential analysis (Squair et al., 2021). Our data lead us to propose that the two radiological phenotypes represent distinct manifestations of a similar pathological process.

T cell infiltration has been a consistent finding in recent studies which have examined the post-COVID-19 airspaces; almost exclusively within 3-6 months of the acute insult, when radiological inflammation is more common than fibrosis (Cheon et al., 2021; Gagiannis et al., 2023; Ravaglia et al., 2022; Vijayakumar et al., 2022). The cellular proportions of our fibrotic PCLD samples, harvested at 9-12 months after acute illness, were akin to those reported in BAL samples from healthy individuals, with macrophages comprising greater than 80% and lymphocytes less than 15% of cells (Mould et al., 2021; The BAL Cooperative Group Steering Committee, 1990). This is in keeping with the reported repopulation of the airspaces by AM in the later stages of COVID-19 acute respiratory distress syndrome (ARDS) (Wendisch et al., 2021). Minimal data currently available for longitudinal samples obtained at approximately one year post-infection also suggest a trajectory of gradual normalization of molecular abnormalities and airspace cellular composition (Vijayakumar et al., 2022). Nonetheless there is considerable interest in therapeutic intervention to expedite resolution of subacute inflammation, to prevent progression to fibrosis with concomitant irreversible tissue damage in susceptible individuals. Our findings suggest therapies which target T cells, rather than anti-fibrotic agents could be beneficial in PCLD, particularly in those with radiological inflammation. In support of this premise, one small uncontrolled clinical study demonstrated improvement in clinical symptoms, physiological parameters and radiological abnormalities, following three weeks of corticosteroid treatment instituted approximately three months post-COVID-19 (Myall et al., 2021).

The bronchoalveolar TCR repertoire from both PCLD phenotypes exhibited high levels of relatedness, suggestive of antigen-specific immune responses. In addition, the high proportion of clusters containing TCRs from multiple donors suggests immune responses against similar antigens, despite almost no sharing of identical TCR sequences between different individuals. Aberrant immune responses to persistent reservoirs of virus have been posited as drivers of post-acute sequelae of SARS-CoV-2 infection (Cheung et al., 2022; Merad et al., 2022; Stein et al., 2022). However, even including all detected TCRs, to offset the inherent sparsity of single-cell data, there was no enrichment for known SARS-CoV-2-specific T cell clones compared to TCRs specific for other common viruses. Hence, based on the current compendium of SARS-CoV-2-reactive TCRs (Bagaev et al., 2020), which may not be comprehensive, there was no evidence for viral persistence at the site of disease in our cohort. A more plausible hypothesis, given reports of cross-reactivity between SARS-CoV-2 and human antigens (Nunez-Castilla et al., 2022; Vojdani and Kharrazian, 2020), is that the PCLD TCR repertoire is directed against as yet unknown respiratory autoantigens. Identifying the antigenic targets of the PCLD T cell repertoire may present opportunities for more specific therapeutic interventions.

Pro-fibrotic macrophages have been implicated in the pathogenesis of severe COVID-19 ARDS, which has been associated with rapid onset pulmonary fibrosis, which improves over time (Wendisch et al., 2021). Minimal interstitial fibrosis and the presence of pro-fibrotic macrophages have also been reported in transbronchial lung biopsies from a subset of individuals sampled at least 12 weeks after mild COVID-19 (Gagiannis et al., 2023). However, there was no difference between the frequency at which mild fibrotic features were detected in the PCLD group and pre-pandemic autopsy samples from individuals who had died of non-respiratory causes (Gagiannis et al., 2023). In our study, which encompassed a broad range of severity of acute disease, including several individuals with COVID-19 ARDS, the phenotype of myeloid cells in both radiological groups was consistent with the spectrum of macrophages and monocytes found in healthy airspaces (Mould et al., 2021); with no convincing evidence for exaggerated activity of pro-fibrogenic pathways. The heterogeneity of disease phenotype and longer time interval after acute illness at which our cohort were sampled may account for this discrepancy. Of note, CMV-specific T cell clones were enriched in individuals with fibrotic PCLD, consistent with the repeated association of CMV with pulmonary fibrosis (Moore and Moore, 2015). However, this observation is of uncertain significance, given the lack of existing evidence for a direct role for CMV in causing human pulmonary fibrosis (Moore and Moore, 2015).

Our study has some limitations. We acknowledge a small sample size, heterogeneous patient cohort, and lack of specimens for histological correlation and orthogonal validation of our single-cell data. Our findings provide early mechanistic insights, are hypothesis-generating and will require further validation in larger cohorts. Those with fibrotic radiological appearances were sampled later after acute disease than those with radiological inflammation. Ideally, we would have more closely matched the interval after acute COVID-19 at which both groups were sampled. However, the introduction of dexamethasone treatment early in the second wave of the pandemic was coincident with reduced numbers of individuals with persistent respiratory symptoms and radiological abnormalities. Consequently, we extended the interval after acute illness within which individuals were eligible for sampling. Since our aim was to evaluate whether single-cell profiling of the bronchoalveolar environment of individuals with inflammatory and fibrotic radiological patterns of PCLD at the time of sampling would reveal different immunopathogenic mechanisms for these phenotypes, we believe our approach remains valid. It also provided invaluable opportunities to assess respiratory tract samples from the later stages of PCLD which have received minimal attention to date. This was a cross-sectional evaluation of bronchoalveolar immune cells, which did not allow assessment of the temporal evolution of the immune response in PCLD within individuals. Future studies encompassing longitudinal monitoring of the bronchoalveolar environment might provide important insights into the molecular mechanisms driving resolution and whether this trajectory is a universal phenomenon for both inflammation and fibrosis following COVID-19. Augmented neutrophil-associated immune signatures have been described in plasma and nasal samples in individuals with post-COVID pulmonary sequelae, however, neutrophils were not detected in our samples, possibly due to the Chromium 10x Genomics sample processing conditions used at the time of our analysis (George et al., 2022). Finally, due to technical limitations because of the low numbers of T cells present in BAL samples, we were unable to obtain TCR data for two inflammatory cases. Nonetheless, our data provide strong support for the notion that the TCR repertoire of PCLD reflects antigen-directed immune responses.

Our observations that inflammatory PCLD is characterised by airway T cell infiltration and that antigen-specific T cell responses are evident in both radiological phenotypes, highlight opportunities for early intervention with therapies targeting T cells. Understanding the timing and duration of intervention, stratification of those at high risk of irreversible tissue damage and the potential use of more targeted T cell immunomodulators all merit further investigation.

## Methods

### Ethics statement

The study was approved by the North London Research Ethics Committee (13/LO/0900). Written informed consent was obtained from all participants. Subject identifiers were not known to anyone outside the research group.

### Study design and eligibility

Immune cells from the site of disease were obtained from adults (≥18 years) undergoing bronchoscopy for clinical investigation of persistent respiratory symptoms and CT abnormalities consistent with pulmonary inflammation (n=5) or fibrosis (n=5) at least 12 weeks after acute COVID-19, confirmed by a positive SARS-CoV-2 antibody or polymerase chain reaction (PCR) test. PCLD was defined in this cohort based on the following criteria: 1) new, persistent respiratory symptoms following SARS-CoV-2 infection at least 12 weeks previously, 2) post-COVID-19 residual lung abnormalities with more than 10% lung involvement on CT and 3) breathlessness in keeping with the CT changes and not explained by other causes. Thoracic CT scans were classified as predominantly inflammatory or fibrotic by consensus opinion of the ILD multi-disciplinary team, which included thoracic radiologists with ILD expertise. Radiological inflammation was defined as consolidation or ground glass opacities without reticulation or parenchymal distortion and fibrosis defined as reticulation or traction bronchiectasis. Individuals with evidence of ILD prior to COVID-19, those with coincident malignancy, human immunodeficiency virus infection, bacterial, viral or fungal respiratory tract infection, taking immunomodulatory therapy, or unable to give informed consent were excluded.

### Isolation of bronchoalveolar cells

Flexible fibreoptic bronchoscopy was used to obtain BAL samples by instillation of 180-240 ml of warmed normal saline into a lung segment affected by the predominant radiological abnormality. Aspirated BAL fluid was cooled to 4°C and filtered through a cell strainer to remove particulate debris before centrifugation. After removal of the supernatant, cells were resuspended in PBS. Cell count and viability were determined by Trypan blue staining and erythrocytes removed where indicated, using ammonium-chloride-potassium red cell lysis buffer. Cells were resuspended at 2 x10^6^ per ml for immediate downstream processing.

### scRNAseq and scTCRseq library preparation and sequencing

20,000 cells per sample were loaded on to the Chromium controller (10x Genomics) to generate single-cell gel beads in emulsion (GEMs). Single-cell partitioning, reverse transcription, cDNA amplification and library construction were performed using the Chromium Single-cell 5’ Reagent kits v1.1 and v2 (10x Genomics) according to the manufacturer’s instructions. T cell receptor (TCR) V(D)J segments were enriched from amplified cDNA using Chromium Single-Cell V(D)J Enrichment kits v1.1 and v2 (10x Genomics) per the manufacturer’s protocol. Libraries were quality checked and quantified using the High Sensitivity DNA kit and 4200 TapeStation (Agilent). Sequencing was performed in paired end mode with SP100, P2 and P3 flow cells (100 cycles) using NovaSeq 6000 and NextSeq 2000 systems (Illumina).

### scRNAseq data analysis

#### Cell Ranger

Raw sequencing files were demultiplexed using BCL Convert v3.7.5 (Illumina) or Cell Ranger version 6.1.1 using the “mkfastq” script. Transcript alignment and quantitation against the GRCh38 human genome assembly was performed using Cell Ranger “multi” for samples with gene expression and T cell VDJ data or Cell Ranger “count” for samples with gene expression data only.

#### Quality control

Initial processing of Cell Ranger output data was performed using Seurat v4.1.0 in R 4.1.1 (Stuart et al., 2019). Cell Ranger output files were loaded using the “Read10x” function. Low quality cells with less than 200 or more than 6000 unique features or more than 10% mitochondrial genes were removed; 57,712 cells were retained for downstream analysis.

#### Normalization, feature selection and integration

For each sample data were normalized and highly variable genes selected before integration to remove donor-specific batch effects, using the following Seurat functions implemented with default parameters; NormalizeData (normalization.method = “LogNormalize), FindVariableFeatures (selection.method = “vst”, nfeatures = 2000), IntegrateData (anchorset = immune.anchors).

#### Dimensional reduction, clustering and annotation

Data were then scaled using the Seurat ScaleData function and dimensional reduction achieved by principal component analysis (PCA) of the most variable genes using RunPCA. The first 25 principal components (PCs) were used to generate the Uniform Manifold Approximation and Projection (UMAP) for two-dimensional visualization of the cells (RunUMAP) and for nearest neighbor graph construction (FindNeighbors) and Louvain clustering (FindClusters) using a resolution of 0.8. This clustering resolution was selected on the basis that it successfully partitioned single-cell transcriptomes into the broad cell types expected to be present in BAL. Automatic cell type annotation of clusters was performed with the Semi-supervised Category Identification and Assignment (SCINA) R package using gene signatures from the Azimuth human lung v1 reference (https://azimuth.hubmapconsortium.org/references/#Human-Lungv1) (Travaglini et al., 2020; Zhang et al., 2019). Additional manual annotation of dendritic cells and B cells was performed using literature-based markers (Cheng et al., 2021; Glass et al., 2020; Villani et al., 2017). A population with low numbers of genes, which could not be annotated by automatic or manual methods and likely represents empty droplets containing ambient RNA, was removed after clustering. To validate our clustering and annotation strategy, we visualized the expression of canonical or published marker genes for each cell type using a dot plot (Adams et al., 2020; Cheng et al., 2021; David et al., 2021; Davies et al., 2013; Davis and Wypych, 2021; Gardell and Parker, 2017; Glass et al., 2020; Murray and Wynn, 2011; Tsyklauri et al., 2023; van Aalderen et al., 2021; Villani et al., 2017).

#### T cell and macrophage re-clustering

For separate analyses of T cells only and macrophages only, cells in the relevant clusters were subsetted and normalization, variable feature selection, integration, data scaling, PCA, UMAP generation and Louvain clustering repeated. The first 25 PCs were used for data integration and clustering for both macrophages and T cells. The k.weight parameter was reduced to 70 for the T cell integration step in order to take account of the low number of cells in sample 12. Resolution 0.4 was used for re-clustering T cells and resolution 0.7 for re-clustering macrophages. T cell subsets were annotated using SCINA with Azimuth human PBMC reference marker genes https://azimuth.hubmapconsortium.org/references/#Human-PBMC and additional published CD4 T cell signatures (Cano-Gamez et al., 2020; Zhang et al., 2019). Macrophages were annotated using SCINA with Azimuth human lung v1 reference marker genes and additional signatures derived from published literature (Mould et al., 2021; Travaglini et al., 2020; Wendisch et al., 2021; Zhang et al., 2019).

#### Calculation of gene module scores

The Seurat function AddModuleScore was used to calculate enrichment of a cyclin D1 (CCND1) regulated module representing the cellular proliferation response (Chandran et al., 2022) and a profibrotic macrophage gene module derived from idiopathic pulmonary fibrosis (Adams et al., 2020). Module scores represent the average expression of gene signatures of interest with subtraction of the average expression of control gene sets randomly selected from each expression bin which contains a module gene.

#### Differential gene expression and differential cell type abundance analysis

For comparison of gene expression in individual cells (single-cell method) the scran R package findMarkers function was implemented using the following settings: test.type = “wilcox”, direction = ‘‘up’’, pval.type = “all”, lfc = 0. For identification of cell type (cluster) marker genes, sample was included as a blocking factor. Genes with an adjusted p-value <0.05 were considered significantly differentially expressed. For comparison of gene expression at cell type level (pseudobulk method) the scuttle R package aggregateAcrossCells function was used to aggregate counts for each cell type within each sample to create “pseudobulks” before statistical analysis was performed. Differentially expressed genes were identified using the scran pseudoBulkDGE function, implementing the edgeR negative binomial generalized linear model with quasi-likelihood F test (GLM-QLF) (Robinson et al., 2010). Genes with an adjusted p-value <0.05 were considered significant. Differences in cell type abundance between inflammatory and fibrotic PCLD were identified using the edgeR GLM-QLF test implemented by scran. Cell types with an adjusted p-value <0.05 were considered differentially abundant.

#### Pathway enrichment analysis

The biological pathways represented by differentially expressed genes were identified by Reactome pathway enrichment analysis using XGR as previously described (Fang et al., 2016; Turner et al., 2021). For visualization, 15 pathway groups were identified by hierarchical clustering of Jaccard indices to quantify similarity between the gene compositions of each pathway. For each group the pathway term with the largest number of annotated genes was then selected as representative of the enriched biology.

#### Upstream regulator analysis

Ingenuity Pathway Analysis (Qiagen) was used to identify upstream transcriptional regulation of differentially expressed genes. This analysis was restricted to molecules annotated with the following functions: cytokine, growth factor, transmembrane receptor, kinase and transcriptional regulator, representing the canonical components of pathways which execute transcriptional reprogramming in immune and tissue repair responses. Enriched molecules with an adjusted p-value <0.05 were considered statistically significant. Area-proportional Venn diagrams visualizing the overlap between molecules predicted to regulate cell type-specific differentially expressed genes in inflammatory PCLD and fibrotic PCLD were generated using BioVenn (Hulsen et al., 2008).

#### TCR quantitation and CDR3 clustering

TCR sequences were assembled by the Cell Ranger multi pipeline (v6.1.1). For single-cell TCR analysis, TCR clonotype abundance information was imported directly from Cell Ranger “filtered_contig_annotations” output files, where clonotype identity was determined as cells with identical V(D)J and CDR3 sequences. Clonotypes were assigned to single cells using index barcodes. TCRs found more than once were defined as expanded. Density plots were calculated using the UMAP coordinates of every expanded cell with a detectable TCR using geom_density_2d from the ggplot2 package.

For clustering analysis of each PCLD phenotype individually, all detected alpha and beta chain sequences in the inflammatory samples were included, to take account of the sparsity of single-cell data. Including larger numbers of TCR sequences leads to more clustering (Madi et al., 2017); fibrotic PCLD and control group TCR sequences were therefore subsampled to match the number of sequences in the inflammatory PCLD group, which contained the smallest repertoire. We included expanded peripheral blood TCRs from individuals with non-severe SARS-CoV-2 infection as a positive control dataset known to exhibit high levels of clustering and non-expanded TCRs randomly selected from uninfected individuals from this same cohort, as a negative control dataset, not expected to cluster highly (Chandran et al., 2022; Milighetti et al., 2023). For the combined analysis of both PCLD phenotypes all detected CDR3 sequences were included. TCR clustering was performed as previously described (Joshi et al., 2019). Briefly, CDR3 amino acid sequences were deconstructed into overlapping series of contiguous triplets. Pairwise similarity between two CDR3s was calculated as the normalized string (triplet) kernel using the Kernlab R package (Karatzoglou et al., 2004). The resulting TCR similarity matrix was converted into a network diagram in which CDR3s with a pairwise similarity above a designated threshold were connected by an edge using the iGraph R package (Csardi and Nepusz, 2006). We visualized thresholds at which PCLD TCRs exhibited the largest increase in the percentage of TCRs clustering compared to negative control TCR data for clustering of PCLD groups individually. For combined clustering of inflammatory and fibrotic PCLD we visualized thresholds at which the largest clusters composed uniquely of one phenotype were present.

#### Virus specific TCR enrichment analysis

TCRs annotated for SARS-CoV-2, CMV and EBV were obtained from the VDJdb database (Bagaev et al., 2020) (https://vdjdb.cdr3.net/), accessed on 1st November 2021. The number of annotated sequences for each virus in VDJdb either matching or not matching TCRs detected in inflammatory and fibrotic PCLD or the negative controls was used to calculate the odds ratio (Fisher’s exact test) for enrichment of virus-specific TCRs in each group.

## Data availability

scRNAseq and scTCRseq data will be available in the Gene Expression Omnibus (GEO) database at the time of peer-reviewed publication of the manuscript under accession number GSE228236.

## Data Availability

scRNAseq and scTCRseq data will be available in the Gene Expression Omnibus (GEO) database at the time of peer-reviewed publication of the manuscript.

## Acknowledgements

We thank Dr Arjun Nair for oversight of the ILD MDT discussions to classify radiological changes as predominantly inflammatory or fibrotic and for identification of lung segments to be targeted for sampling by BAL.

## Author contributions

GST, JCP, RCC and MZN conceived the study.

EKD, PM, KBW, MY, JCP and GST collected the data.

PM, EKD, KW, MY 10x Genomics platform.

BSMDDE, CTT, KF, MM, MZN, MP, JSB, BMC and MN provided resources or analysis tools.

BSMDDE, KF and GST performed the computational data analysis with contributions from AM, MM, BMC and MN.

PM, BSMDDE, EKD and GST prepared the manuscript draft.

All authors reviewed and contributed to the final manuscript.

## References

Adams TS, Schupp JC, Poli S, Ayaub EA, Neumark N, Ahangari F, Chu SG, Raby BA, DeIuliis G, Januszyk M, Duan Q, Arnett HA, Siddiqui A, Washko GR, Homer R, Yan X, Rosas IO, Kaminski N. 2020. Single-cell RNA-seq reveals ectopic and aberrant lung-resident cell populations in idiopathic pulmonary fibrosis. Sci Adv 6:eaba1983. doi:10.1126/sciadv.aba1983

Auffray C, Sieweke MH, Geissmann F. 2009. Blood monocytes: development, heterogeneity, and relationship with dendritic cells. Annu Rev Immunol 27:669–692. doi:10.1146/annurev.immunol.021908.132557

Bagaev DV, Vroomans RMA, Samir J, Stervbo U, Rius C, Dolton G, Greenshields-Watson A, Attaf M, Egorov ES, Zvyagin IV, Babel N, Cole DK, Godkin AJ, Sewell AK, Kesmir C, Chudakov DM, Luciani F, Shugay M. 2020. VDJdb in 2019: database extension, new analysis infrastructure and a T-cell receptor motif compendium. Nucleic Acids Res 48:D1057–D1062. doi:10.1093/nar/gkz874

Cano-Gamez E, Soskic B, Roumeliotis TI, So E, Smyth DJ, Baldrighi M, Willé D, Nakic N, Esparza-Gordillo J, Larminie CGC, Bronson PG, Tough DF, Rowan WC, Choudhary JS, Trynka G. 2020. Single-cell transcriptomics identifies an effectorness gradient shaping the response of CD4+ T cells to cytokines. Nat Commun 11:1801. doi:10.1038/s41467-020-15543-y

Chandran A, Rosenheim J, Nageswaran G, Swadling L, Pollara G, Gupta RK, Burton AR, Guerra-Assunção JA, Woolston A, Ronel T, Pade C, Gibbons JM, Sanz-Magallon Duque De Estrada B, Robert de Massy M, Whelan M, Semper A, Brooks T, Altmann DM, Boyton RJ, McKnight Á, Captur G, Manisty C, Treibel TA, Moon JC, Tomlinson GS, Maini MK, Chain BM, Noursadeghi M, COVIDsortium Investigators. 2022. Rapid synchronous type 1 IFN and virus-specific T cell responses characterize first wave non-severe SARS-CoV-2 infections. Cell Rep Med 3:100557. doi:10.1016/j.xcrm.2022.100557

Cheng S, Li Z, Gao R, Xing B, Gao Y, Yang Yu, Qin S, Zhang L, Ouyang H, Du P, Jiang L, Zhang B, Yang Yue, Wang X, Ren X, Bei J-X, Hu X, Bu Z, Ji J, Zhang Z. 2021. A pan-cancer single-cell transcriptional atlas of tumor infiltrating myeloid cells. Cell 184:792–809.e23. doi:10.1016/j.cell.2021.01.010

Cheon IS, Li C, Son YM, Goplen NP, Wu Y, Cassmann T, Wang Z, Wei X, Tang J, Li Y, Marlow H, Hughes S, Hammel L, Cox TM, Goddery E, Ayasoufi K, Weiskopf D, Boonyaratanakornkit J, Dong H, Li H, Chakraborty R, Johnson AJ, Edell E, Taylor JJ, Kaplan MH, Sette A, Bartholmai BJ, Kern R, Vassallo R, Sun J. 2021. Immune signatures underlying post-acute COVID-19 lung sequelae. Sci Immunol 6:eabk1741. doi:10.1126/sciimmunol.abk1741

Cheung CCL, Goh D, Lim X, Tien TZ, Lim JCT, Lee JN, Tan B, Tay ZEA, Wan WY, Chen EX, Nerurkar SN, Loong S, Cheow PC, Chan CY, Koh YX, Tan TT, Kalimuddin S, Tai WMD, Ng JL, Low JG-H, Yeong J, Lim KH. 2022. Residual SARS-CoV-2 viral antigens detected in GI and hepatic tissues from five recovered patients with COVID-19. Gut 71:226–229. doi:10.1136/gutjnl-2021-324280

Csardi G, Nepusz T. 2006. The igraph software package for complex network research. Complex Syst 1695:1–9.

David G, Willem C, Legrand N, Djaoud Z, Mérieau P, Walencik A, Guillaume T, Gagne K, Chevallier P, Retière C. 2021. Deciphering the biology of KIR2DL3+ T lymphocytes that are associated to relapse in haploidentical HSCT. Sci Rep 11:15782. doi:10.1038/s41598-021-95245-7

Davies LC, Jenkins SJ, Allen JE, Taylor PR. 2013. Tissue-resident macrophages. Nat Immunol 14:986–995. doi:10.1038/ni.2705

Davis JD, Wypych TP. 2021. Cellular and functional heterogeneity of the airway epithelium. Mucosal Immunol 14:978–990. doi:10.1038/s41385-020-00370-7

Fabbri L, Moss S, Khan FA, Chi W, Xia J, Robinson K, Smyth AR, Jenkins G, Stewart I. 2023. Parenchymal lung abnormalities following hospitalisation for COVID-19 and viral pneumonitis: a systematic review and meta-analysis. Thorax 78:191–201. doi:10.1136/thoraxjnl-2021-218275

Fang H, Knezevic B, Burnham KL, Knight JC. 2016. XGR software for enhanced interpretation of genomic summary data, illustrated by application to immunological traits. Genome Med 8:129. doi:10.1186/s13073-016-0384-y

Gagiannis D, Hackenbroch C, Bloch W, Zech F, Kirchhoff F, Djudjaj S, von Stillfried S, Bülow R, Boor P, Steinestel K. 2023. Clinical, Imaging, and Histopathological Features of Pulmonary Sequelae Following Mild COVID-19. Am J Respir Crit Care Med. doi:10.1164/rccm.202302-0285LE

Gardell JL, Parker DC. 2017. CD40L is transferred to antigen-presenting B cells during delivery of T-cell help. Eur J Immunol 47:41–50. doi:10.1002/eji.201646504

George PM, Reed A, Desai SR, Devaraj A, Faiez TS, Laverty S, Kanwal A, Esneau C, Liu MKC, Kamal F, Man WD-C, Kaul S, Singh S, Lamb G, Faizi FK, Schuliga M, Read J, Burgoyne T, Pinto AL, Micallef J, Bauwens E, Candiracci J, Bougoussa M, Herzog M, Raman L, Ahmetaj-Shala B, Turville S, Aggarwal A, Farne HA, Dalla Pria A, Aswani AD, Patella F, Borek WE, Mitchell JA, Bartlett NW, Dokal A, Xu X-N, Kelleher P, Shah A, Singanayagam A. 2022. A persistent neutrophil-associated immune signature characterizes post-COVID-19 pulmonary sequelae. Sci Transl Med 14:eabo5795. doi:10.1126/scitranslmed.abo5795

Glass DR, Tsai AG, Oliveria JP, Hartmann FJ, Kimmey SC, Calderon AA, Borges L, Glass MC, Wagar LE, Davis MM, Bendall SC. 2020. An Integrated Multi-omic Single-Cell Atlas of Human B Cell Identity. Immunity 53:217–232.e5. doi:10.1016/j.immuni.2020.06.013

Hulsen T, de Vlieg J, Alkema W. 2008. BioVenn - a web application for the comparison and visualization of biological lists using area-proportional Venn diagrams. BMC Genomics 9:488. doi:10.1186/1471-2164-9-488

Joshi K, de Massy MR, Ismail M, Reading JL, Uddin I, Woolston A, Hatipoglu E, Oakes T, Rosenthal R, Peacock T, Ronel T, Noursadeghi M, Turati V, Furness AJS, Georgiou A, Wong YNS, Ben Aissa A, Sunderland MW, Jamal-Hanjani M, Veeriah S, Birkbak NJ, Wilson GA, Hiley CT, Ghorani E, Guerra-Assunção JA, Herrero J, Enver T, Hadrup SR, Hackshaw A, Peggs KS, McGranahan N, Swanton C, TRACERx consortium, Quezada SA, Chain B. 2019. Spatial heterogeneity of the T cell receptor repertoire reflects the mutational landscape in lung cancer. Nat Med 25:1549–1559. doi:10.1038/s41591-019-0592-2

Karatzoglou A, Smola A, Hornik K, Zeileis A. 2004. kernlab - An S4 Package for Kernel Methods in R. J Stat Softw 11:1–20. doi:10.18637/jss.v011.i09

Liao M, Liu Y, Yuan J, Wen Y, Xu G, Zhao J, Cheng L, Li J, Wang X, Wang F, Liu L, Amit I, Zhang S, Zhang Z. 2020. Single-cell landscape of bronchoalveolar immune cells in patients with COVID-19. Nat Med 26:842–844. doi:10.1038/s41591-020-0901-9

Madi A, Poran A, Shifrut E, Reich-Zeliger S, Greenstein E, Zaretsky I, Arnon T, Laethem FV, Singer A, Lu J, Sun PD, Cohen IR, Friedman N. 2017. T cell receptor repertoires of mice and humans are clustered in similarity networks around conserved public CDR3 sequences. eLife 6:e22057. doi:10.7554/eLife.22057

Mehta P, Rosas IO, Singer M. 2022. Understanding post-COVID-19 interstitial lung disease (ILD): a new fibroinflammatory disease entity. Intensive Care Med 48:1803–1806. doi:10.1007/s00134-022-06877-w

Merad M, Blish CA, Sallusto F, Iwasaki A. 2022. The immunology and immunopathology of COVID-19. Science 375:1122–1127. doi:10.1126/science.abm8108

Milighetti M, Peng Y, Tan C, Mark M, Nageswaran G, Byrne S, Ronel T, Peacock T, Mayer A, Chandran A, Rosenheim J, Whelan M, Yao X, Liu G, Felce SL, Dong T, Mentzer AJ, Knight JC, Balloux F, Greenstein E, Reich-Zeliger S, Pade C, Gibbons JM, Semper A, Brooks T, Otter A, Altmann DM, Boyton RJ, Maini MK, McKnight A, Manisty C, Treibel TA, Moon JC, Noursadeghi M, Chain B. 2023. Large clones of pre-existing T cells drive early immunity against SARS-COV-2 and LCMV infection. iScience 26. doi:10.1016/j.isci.2023.106937

Moore BB, Moore TA. 2015. Viruses in Idiopathic Pulmonary Fibrosis. Etiology and Exacerbation. Ann Am Thorac Soc 12 Suppl 2:S186–192. doi:10.1513/AnnalsATS.201502-088AW

Mould KJ, Moore CM, McManus SA, McCubbrey AL, McClendon JD, Griesmer CL, Henson PM, Janssen WJ. 2021. Airspace Macrophages and Monocytes Exist in Transcriptionally Distinct Subsets in Healthy Adults. Am J Respir Crit Care Med 203:946–956. doi:10.1164/rccm.202005-1989OC

Murray PJ, Wynn TA. 2011. Protective and pathogenic functions of macrophage subsets. Nat Rev Immunol 11:723–737. doi:10.1038/nri3073

Myall KJ, Mukherjee B, Castanheira AM, Lam JL, Benedetti G, Mak SM, Preston R, Thillai M, Dewar A, Molyneaux PL, West AG. 2021. Persistent Post-COVID-19 Interstitial Lung Disease. An Observational Study of Corticosteroid Treatment. Ann Am Thorac Soc 18:799–806. doi:10.1513/AnnalsATS.202008-1002OC

Nouno T, Okamoto M, Ohnishi K, Kaieda S, Tominaga M, Zaizen Y, Ichiki M, Momosaki S, Nakamura M, Fujimoto K, Fukuoka J, Shimizu S, Komohara Y, Hoshino T. 2019. Elevation of pulmonary CD163 + and CD204 + macrophages is associated with the clinical course of idiopathic pulmonary fibrosis patients. J Thorac Dis 11. doi:10.21037/jtd.2019.09.03

Nunez-Castilla J, Stebliankin V, Baral P, Balbin CA, Sobhan M, Cickovski T, Mondal AM, Narasimhan G, Chapagain P, Mathee K, Siltberg-Liberles J. 2022. Potential Autoimmunity Resulting from Molecular Mimicry between SARS-CoV-2 Spike and Human Proteins. Viruses 14:1415. doi:10.3390/v14071415

Ravaglia C, Doglioni C, Chilosi M, Piciucchi S, Dubini A, Rossi G, Pedica F, Puglisi S, Donati L, Tomassetti S, Poletti V. 2022. Clinical, radiological and pathological findings in patients with persistent lung disease following SARS-CoV-2 infection. Eur Respir J 60:2102411. doi:10.1183/13993003.02411-2021

Robinson MD, McCarthy DJ, Smyth GK. 2010. edgeR: a Bioconductor package for differential expression analysis of digital gene expression data. Bioinforma Oxf Engl 26:139–140. doi:10.1093/bioinformatics/btp616

Squair JW, Gautier M, Kathe C, Anderson MA, James ND, Hutson TH, Hudelle R, Qaiser T, Matson KJE, Barraud Q, Levine AJ, La Manno G, Skinnider MA, Courtine G. 2021. Confronting false discoveries in single-cell differential expression. Nat Commun 12:5692. doi:10.1038/s41467-021-25960-2

Stein SR, Ramelli SC, Grazioli A, Chung J-Y, Singh M, Yinda CK, Winkler CW, Sun J, Dickey JM, Ylaya K, Ko SH, Platt AP, Burbelo PD, Quezado M, Pittaluga S, Purcell M, Munster VJ, Belinky F, Ramos-Benitez MJ, Boritz EA, Lach IA, Herr DL, Rabin J, Saharia KK, Madathil RJ, Tabatabai A, Soherwardi S, McCurdy MT, NIH COVID-19 Autopsy Consortium, Peterson KE, Cohen JI, de Wit E, Vannella KM, Hewitt SM, Kleiner DE, Chertow DS. 2022. SARS-CoV-2 infection and persistence in the human body and brain at autopsy. Nature 612:758–763. doi:10.1038/s41586-022-05542-y

Stewart I, Jacob J, George PM, Molyneaux PL, Porter JC, Allen RJ, Aslani S, Baillie JK, Barratt SL, Beirne P, Bianchi SM, Blaikley JF, Chalmers JD, Chambers RC, Chaudhuri N, Coleman C, Collier G, Denneny EK, Docherty A, Elneima O, Evans RA, Fabbri L, Gibbons MA, Gleeson FV, Gooptu B, Greening NJ, Guillen Guio B, Hall IP, Hanley NA, Harris V, Harrison EM, Heightman M, Hillman TE, Horsley A, Houchen-Wolloff L, Jarrold I, Johnson SR, Jones MG, Khan F, Lawson R, Leavy O, Lone N, Marks M, McAuley H, Mehta P, Parekh D, Piper Hanley K, Platé M, Pearl J, Poinasamy K, Quint JK, Raman B, Richardson M, Rivera-Ortega P, Saunders L, Saunders R, Semple MG, Sereno M, Shikotra A, Simpson AJ, Singapuri A, Smith DJ, Spears M, Spencer LG, Stanel S, Thickett D, Thompson AAR, Thorpe M, Walsh SL, Walker S, Weatherley ND, Weeks M, Wild JM, Wootton DG, Brightling CE, Ho L-P, Wain LV, Jenkins RG. 2022. Residual Lung Abnormalities Following COVID-19 Hospitalization: Interim Analysis of the UKILD Post-COVID Study. Am J Respir Crit Care Med. doi:10.1164/rccm.202203-0564OC

Stuart T, Butler A, Hoffman P, Hafemeister C, Papalexi E, Mauck WM, Hao Y, Stoeckius M, Smibert P, Satija R. 2019. Comprehensive Integration of Single-Cell Data. Cell 177:1888–1902.e21. doi:10.1016/j.cell.2019.05.031

The BAL Cooperative Group Steering Committee. 1990. Bronchoalveolar lavage constituents in healthy individuals, idiopathic pulmonary fibrosis, and selected comparison groups. Am Rev Respir Dis 141:S169–202. doi:10.1164/ajrccm/141.5_Pt_2.S169

Travaglini KJ, Nabhan AN, Penland L, Sinha R, Gillich A, Sit RV, Chang S, Conley SD, Mori Y, Seita J, Berry GJ, Shrager JB, Metzger RJ, Kuo CS, Neff N, Weissman IL, Quake SR, Krasnow MA. 2020. A molecular cell atlas of the human lung from single-cell RNA sequencing. Nature 587:619–625. doi:10.1038/s41586-020-2922-4

Tsyklauri O, Chadimova T, Niederlova V, Kovarova J, Michalik J, Malatova I, Janusova S, Ivashchenko O, Rossez H, Drobek A, Vecerova H, Galati V, Kovar M, Stepanek O. 2023. Regulatory T cells suppress the formation of potent KLRK1 and IL-7R expressing effector CD8 T cells by limiting IL-2. eLife 12:e79342. doi:10.7554/eLife.79342

Turner CT, Brown J, Shaw E, Uddin I, Tsaliki E, Roe JK, Pollara G, Sun Y, Heather JM, Lipman M, Chain B, Noursadeghi M. 2021. Persistent T Cell Repertoire Perturbation and T Cell Activation in HIV After Long Term Treatment. Front Immunol 12:634489. doi:10.3389/fimmu.2021.634489

van Aalderen MC, van Lier RAW, Hombrink P. 2021. How to Reliably Define Human CD8+ T-Cell Subsets: Markers Playing Tricks. Cold Spring Harb Perspect Biol 13:a037747. doi:10.1101/cshperspect.a037747

Vijayakumar B, Boustani K, Ogger PP, Papadaki A, Tonkin J, Orton CM, Ghai P, Suveizdyte K, Hewitt RJ, Desai SR, Devaraj A, Snelgrove RJ, Molyneaux PL, Garner JL, Peters JE, Shah PL, Lloyd CM, Harker JA. 2022. Immuno-proteomic profiling reveals aberrant immune cell regulation in the airways of individuals with ongoing post-COVID-19 respiratory disease. Immunity 55:542–556.e5. doi:10.1016/j.immuni.2022.01.017

Villani A-C, Satija R, Reynolds G, Sarkizova S, Shekhar K, Fletcher J, Griesbeck M, Butler A, Zheng S, Lazo S, Jardine L, Dixon D, Stephenson E, Nilsson E, Grundberg I, McDonald D, Filby A, Li W, De Jager PL, Rozenblatt-Rosen O, Lane AA, Haniffa M, Regev A, Hacohen N. 2017. Single-cell RNA-seq reveals new types of human blood dendritic cells, monocytes, and progenitors. Science 356:eaah4573. doi:10.1126/science.aah4573

Vojdani A, Kharrazian D. 2020. Potential antigenic cross-reactivity between SARS-CoV-2 and human tissue with a possible link to an increase in autoimmune diseases. Clin Immunol 217:108480. doi:10.1016/j.clim.2020.108480

Wauters E, Van Mol P, Garg AD, Jansen S, Van Herck Y, Vanderbeke L, Bassez A, Boeckx B, Malengier-Devlies B, Timmerman A, Van Brussel T, Van Buyten T, Schepers R, Heylen E, Dauwe D, Dooms C, Gunst J, Hermans G, Meersseman P, Testelmans D, Yserbyt J, Tejpar S, De Wever W, Matthys P, Neyts J, Wauters J, Qian J, Lambrechts D. 2021. Discriminating mild from critical COVID-19 by innate and adaptive immune single-cell profiling of bronchoalveolar lavages. Cell Res 31:272–290. doi:10.1038/s41422-020-00455-9

Wendisch D, Dietrich O, Mari T, von Stillfried S, Ibarra IL, Mittermaier M, Mache C, Chua RL, Knoll R, Timm S, Brumhard S, Krammer T, Zauber H, Hiller AL, Pascual-Reguant A, Mothes R, Bülow RD, Schulze J, Leipold AM, Djudjaj S, Erhard F, Geffers R, Pott F, Kazmierski J, Radke J, Pergantis P, Baßler K, Conrad Claudia, Aschenbrenner AC, Sawitzki B, Landthaler M, Wyler E, Horst D, Deutsche COVID-19 OMICS Initiative (DeCOI), Hippenstiel S, Hocke A, Heppner FL, Uhrig A, Garcia C, Machleidt F, Herold S, Elezkurtaj S, Thibeault C, Witzenrath M, Cochain C, Suttorp N, Drosten C, Goffinet C, Kurth F, Schultze JL, Radbruch H, Ochs M, Eils R, Müller-Redetzky H, Hauser AE, Luecken MD, Theis FJ, Conrad Christian, Wolff T, Boor P, Selbach M, Saliba A-E, Sander LE. 2021. SARS-CoV-2 infection triggers profibrotic macrophage responses and lung fibrosis. Cell 184:6243–6261.e27. doi:10.1016/j.cell.2021.11.033

Zhang Z, Luo D, Zhong X, Choi JH, Ma Y, Wang S, Mahrt E, Guo W, Stawiski EW, Modrusan Z, Seshagiri S, Kapur P, Hon GC, Brugarolas J, Wang T. 2019. SCINA: A Semi-Supervised Subtyping Algorithm of Single Cells and Bulk Samples. Genes 10:531. doi:10.3390/genes10070531

