## Supplementary information for "Single-cell analysis of bronchoalveolar cells in inflammatory and fibrotic post-COVID lung disease"

### SUPPLEMENTARY FIGURE LEGENDS

**Supplementary Figure 1.** Cellular composition of post-COVID-19 lung disease (PCLD) bronchoalveolar lavage (BAL) samples (related to Figure 1). **a-c** Uniform manifold approximation and projection (UMAP) embedding of 55,776 PCLD BAL single-cell transcriptomes color coded by **a** cell type **b** donor and **c** radiological phenotype. **d** Heatmap of up to the top 50 differentially expressed genes (Wilcoxon test, FDR<0.05) for each cell type and across subjects, colored by z-scores of log-normalized mRNA counts.

**Supplementary Figure 2.** Biological pathways enriched within cell type-specific differentially expressed genes in each post-COVID-19 lung disease (PCLD) phenotype (related to Figure 1). Enrichment of Reactome pathways for cell type-specific genes expressed at significantly higher levels in **a** inflammatory PCLD and **b** fibrotic PCLD, identified by Wilcoxon test (FDR<0.05). Dot size represents the number of genes overlapping each biological pathway and colors reflect z-scores as an indicator of statistical significance.

**Supplementary Figure 3.** Predicted cytokine and growth factor upstream regulators of cell type-specific differential gene expression in post-COVID-19 lung disease (PCLD) (related to Figure 1). Heatmaps display integrated lists of the top 10 most statistically significant cytokines (orange) and growth factors (blue) across all cell types, predicted to regulate cell type-specific differentially expressed genes expressed more highly in **a,d** inflammatory PCLD and **b,e** fibrotic PCLD. Colors indicate statistical significance, represented by -log10FDR values. Grey heatmap cells indicate instances where molecules were not predicted to be upstream regulators of differential gene expression in that particular cell type. Area-proportional Venn diagrams represent the overlap of **c** cytokines and **f** growth factors predicted to regulate cell type-specific differential gene expression in inflammatory and fibrotic PCLD.

**Supplementary Figure 4.** Enriched biological pathways among bronchoalveolar T cell subset-specific differentially expressed genes in post-COVID-19 lung disease (PCLD) (related to Figure 2). Enrichment of Reactome pathways for genes expressed at significantly higher levels in T cell subsets in **a** inflammatory PCLD and **b** fibrotic PCLD, identified by Wilcoxon test (FDR<0.05). Dot size represents the number of genes overlapping each biological pathway and colors represent statistical significance defined by z-score.

**Supplementary Figure 5.** Predicted upstream regulation of T cell subset-specific differential gene expression in inflammatory and fibrotic post-COVID-19 lung disease (PCLD) (related to Figure 2). Heatmap visualisation of integrated lists of the top 10 most statistically enriched cytokines predicted to regulate differentially expressed genes specific to each of the three major T cell subsets expressed at higher levels in **a** inflammatory PCLD and **b** fibrotic PCLD. Colors indicate statistical significance, represented by -log10FDR values. Grey heatmap cells represent molecules not predicted to be upstream regulators of differential gene expression in a particular T cell subset. **c** Area-proportional Venn diagram representing the overlap of cytokines predicted to regulate T cell subset-specific differential gene expression in inflammatory and fibrotic PCLD.

**Supplementary Figure 6.** Macrophage marker gene expression and enriched biological pathways in post-COVID-19 lung disease (PCLD) myeloid populations (related to Figure 3). **a** Dot plot displays expression levels of independently established macrophage marker genes in airspace macrophage and monocyte subsets identified in PCLD. Dot size indicates the percentage of cells expressing the gene in each population, color represents average expression of scaled, log-normalized mRNA counts. **b** Enrichment of Reactome pathways for differentially expressed genes expressed more highly in each myeloid subset, identified by Wilcoxon test (FDR<0.05). Dot size represents the number of genes overlapping each biological pathway and colors represent statistical significance measured by z-score.

**Supplementary Figure 7.** Biological pathways enriched among myeloid subset-specific differentially expressed genes in each post-COVID-19 lung disease (PCLD) phenotype (related to Figure 3). **a** Expression of a profibrotic macrophage gene signature derived from idiopathic pulmonary fibrosis, calculated on a single-cell level, colored by module score and projected on to the macrophage UMAP which is split by radiological phenotype. **b,c** Enrichment of Reactome pathways for genes expressed at significantly higher levels in each macrophage and monocyte subset in **b** inflammatory PCLD and **c** fibrotic PCLD, identified by Wilcoxon test (FDR<0.05). Dot size represents the number of genes overlapping each biological pathway and colors reflect z-scores as an indicator of statistical significance.

**Supplementary Figure 8.** Predicted upstream regulation of differential gene expression in macrophage and monocyte subsets in inflammatory and fibrotic post-COVID-19 lung disease (PCLD) (related to Figure 3). Heatmaps depict integrated lists of the top 10 most statistically enriched cytokines (orange) or growth factors (blue) predicted to regulate differentially expressed genes expressed at higher levels each myeloid population in **a,d** inflammatory PCLD and **b,e** fibrotic PCLD. Colors indicate statistical significance, defined by -log10FDR values. Molecules not predicted to be upstream regulators of differential gene expression in a particular subset are displayed as grey heatmap cells. Area-proportional Venn diagrams represent the overlap of **c** cytokines and **f** growth factors predicted to regulate myeloid subset-specific differential gene expression in inflammatory and fibrotic PCLD.

### SUPPLEMENTARY TABLES

**Supplementary Table 1.** Clinical and demographic information.

| Subject ID | CT | Age range | Sex | Ethnicity | BMI | Smoking status | Steroid | Immune | Respiratory support | BAL‖‖ (days post acute-COVID) |
|---|---|---|---|---|---|---|---|---|---|---|
| 1 | Inf* | 61-65 | Female | Asian | 23.9 | Never | No | No | I&V** | 97 |
| 3 | Inf | 46-50 | Female | Asian | 31.8 | Never | MP‡ | Tocilizumab | I&V, ECMO†† | 159 |
| 4 | Fib† | 51-55 | Male | Asian | 27.3 | Ex | Dex§ | No | HFNO‡‡ | 336 |
| 5 | Fib | 61-65 | Male | White | 23.2 | Ex | Dex | No | I&V | 314 |
| 6 | Fib | 56-60 | Male | White | 30.6 | Never | Dex | No | I&V | 351 |
| 7 | Fib | 61-65 | Male | Asian | 27.4 | Never | MP, Pred‖ | No | I&V | 316 |
| 8 | Fib | 56-60 | Female | White | 22.6 | Ex | Pred | Tocilizumab | I&V | 272 |
| 9 | Inf | 41-45 | Male | White | 37.3 | Never | No | No | Nil | 111 |
| 12 | Inf | 66-70 | Female | White | 46.5 | Never | Dex | No | CPAP§§ | 149 |
| 13 | Inf | 61-65 | Male | White | 28.7 | Ex | Dex | No | HFNO | 116 |

Clinical and demographic data are provided for all study subjects. *Inf = Inflammatory, †Fib = fibrotic, ‡MP = methylprednisolone, §Dex = dexamethasone, ‖Pred = prednisolone, **I&V = intubation and ventilation, ††ECMO = extracorporeal membrane oxygenation, ‡‡HFNO = high flow nasal oxygen, §§CPAP = continuous positive airway pressure, ‖‖BAL = bronchoalveolar lavage.

**Supplementary Table 2.** Cell type-specific differentially expressed genes identified in pseudobulk data.

| **Full_dataset_up_inf** | | | |
|---|---|---|---|
| logFC | FDR | cluster | gene |
| 2.010314 | 0.025086 | Dendritic | *RHOB* |
| 2.918473 | 4.39E-06 | Prolif | *SKAP1* |
| 2.241937 | 0.001254 | Prolif | *CD3D* |
| 1.637594 | 0.002353 | Prolif | *RHOB* |
| 1.944616 | 0.002504 | Prolif | *CLEC2D* |
| 1.961607 | 0.002504 | Prolif | *CD3G* |
| 2.293015 | 0.002504 | Prolif | *FOS* |
| 1.601146 | 0.002504 | Prolif | *JUNB* |
| 2.097024 | 0.005734 | Prolif | *PHLDA1* |
| 1.478716 | 0.013705 | Prolif | *ATP2B1-AS1* |
| 2.55377 | 0.013705 | Prolif | *IFITM1* |
| 1.679621 | 0.032128 | Prolif | *PTPN7* |
| 1.317954 | 0.046678 | Prolif | *ICAM3* |
| **Full_dataset_up_fib** | | | |
| logFC | FDR | cluster | gene |
| 1.654557 | 0.002084 | Prolif | *SAP30* |
| **Tcell_up_inf** | | | |
| logFC | FDR | cluster | gene |
| **Tcell_up_fib** | | | |
| logFC | FDR | cluster | gene |
| **Myeloid_up_inf** | | | |
| logFC | FDR | cluster | gene |
| 2.040356 | 0.010432 | *FCN1*-Mono | *GADD45B* |
| 2.095153 | 0.026693 | *FCN1*-Mono | *RGS2* |
| 1.453707 | 0.046217 | *FCN1*-Mono | *LGMN* |
| 2.154559 | 9.43E-05 | IFN stim AM | *FOS* |
| 1.651666 | 0.001844 | IFN stim AM | *JUNB* |
| 1.710627 | 0.002371 | IFN stim AM | *LGMN* |
| 1.698431 | 0.002683 | IFN stim AM | *DUSP1* |
| 1.309848 | 0.020696 | IFN stim AM | *RHOB* |
| 1.869253 | 4.90E-05 | MT-AM | *DUSP1* |
| 1.845162 | 0.000164 | MT-AM | *FOS* |
| 1.563411 | 0.000164 | MT-AM | *JUNB* |
| 1.236468 | 0.011581 | MT-AM | *RHOB* |
| 1.582065 | 0.04207 | MT-AM | *LGMN* |

| | | | |
|---|---|---|---|
| 1.918576 | 0.036913 | Prolif AM | *RHOB* |
| **Myeloid_up_fib** | | | |
| logFC | FDR | cluster | gene |
| 1.695279 | 0.048914 | *FCN1*-Mono | *ZFPM1* |
| 1.616057 | 0.002683 | IFN stim AM | *RRAS* |
| 1.33114 | 0.01932 | IFN stim AM | *CA2* |
| 1.765763 | 0.01323 | MT-AM | *UQCRHL* |
| 1.682482 | 0.020917 | MT-AM | *RETN* |
| 1.416307 | 0.024438 | MT-AM | *MT1M* |
| 1.489704 | 0.042579 | MT-AM | *CCL23* |

The results of differential expression analysis using a negative binomial generalized linear model with quasi-likelihood F test (GLM-QLF) performed on data aggregated to cell type pseudobulk level for each donor for the full dataset, T cells only and myeloid cells only. A false discovery rate (FDR) <0.05 was considered significant. “Up_inf” denotes genes expressed at significantly higher level in inflammatory PCLD and “up_fib” denotes genes expressed at significantly higher level in fibrotic PCLD. logFC = log2 fold change comparing inflammatory and fibrotic PCLD, cluster indicates cell type. Empty cells indicate that no genes were identified as differentially expressed between the two radiological groups in any T cell subset.

**Supplementary Table 3**. Shared CDR3 sequences.

| Sample | CT | CDR3 | Chain | Frequency |
|---|---|---|---|---|
| 9 | Inflammatory | CAVNTNAGKSTF | Alpha | 1 |
| 13 | Inflammatory | CAVNTNAGKSTF | Alpha | 2 |
| 4 | Fibrotic | CAVRDSNYQLIW | Alpha | 1 |
| 6 | Fibrotic | CAVRDSNYQLIW | Alpha | 1 |
| 7 | Fibrotic | CAVRDSNYQLIW | Alpha | 1 |
| 5 | Fibrotic | CAVRPRSGNTPLVF | Alpha | 1 |
| 7 | Fibrotic | CAVRPRSGNTPLVF | Alpha | 1 |

CDR3 amino acid sequences found in more than one subject are listed, along with the frequency at which they were detected.

**Supplementary Table 4.** Virus-reactive CDR3 sequences in clusters composed of one PCLD phenotype.

| Sample | CT | Cluster | CDR3 sequence | Chain | Virus |
|---|---|---|---|---|---|
| 7 | Fib* | 82 | CAVNTGFQKLVF | Alpha | SARS-CoV-2‡ |
| 13 | Inf† | 77 | CAVGAGTNAGKSTF | Alpha | CMV§ |

List of virus-reactive CDR3 amino acid sequences from VDJdb detected in clusters composed uniquely of one PCLD phenotype. *Fib = Fibrotic, †Inf = inflammatory, ‡SARS-CoV-2 = severe acute respiratory syndrome coronavirus 2, §CMV = cytomegalovirus.

Supplementary Figure 1

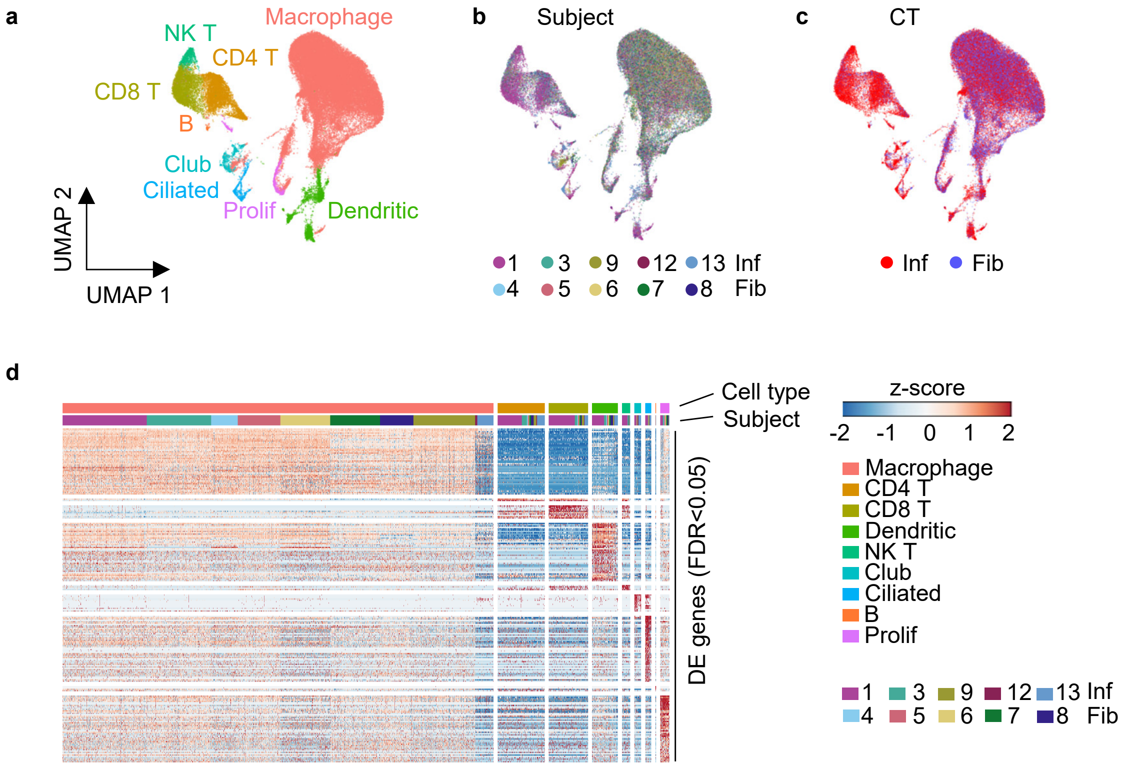

Supplementary Figure 2

**a**

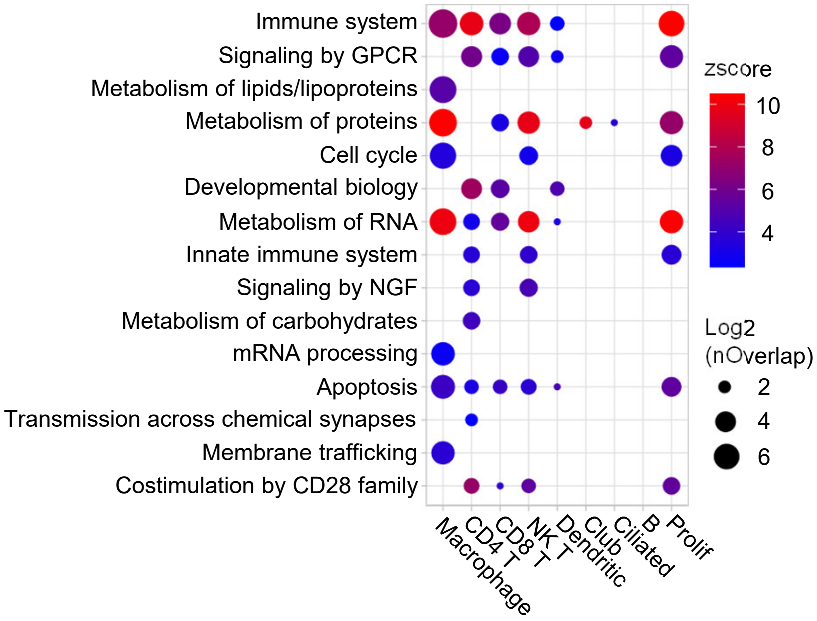

**b**

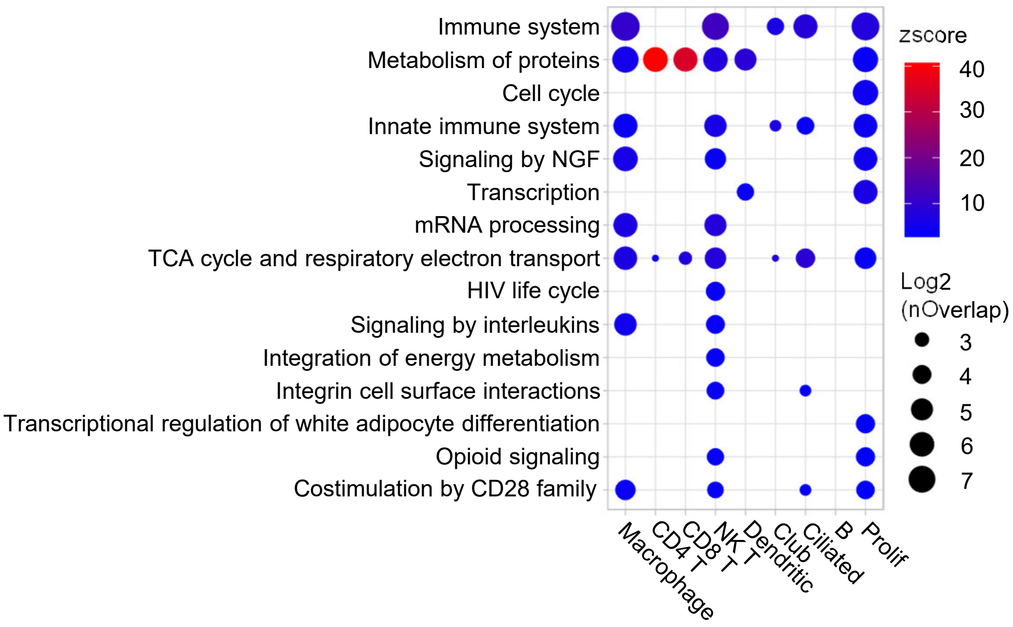

Supplementary Figure 3

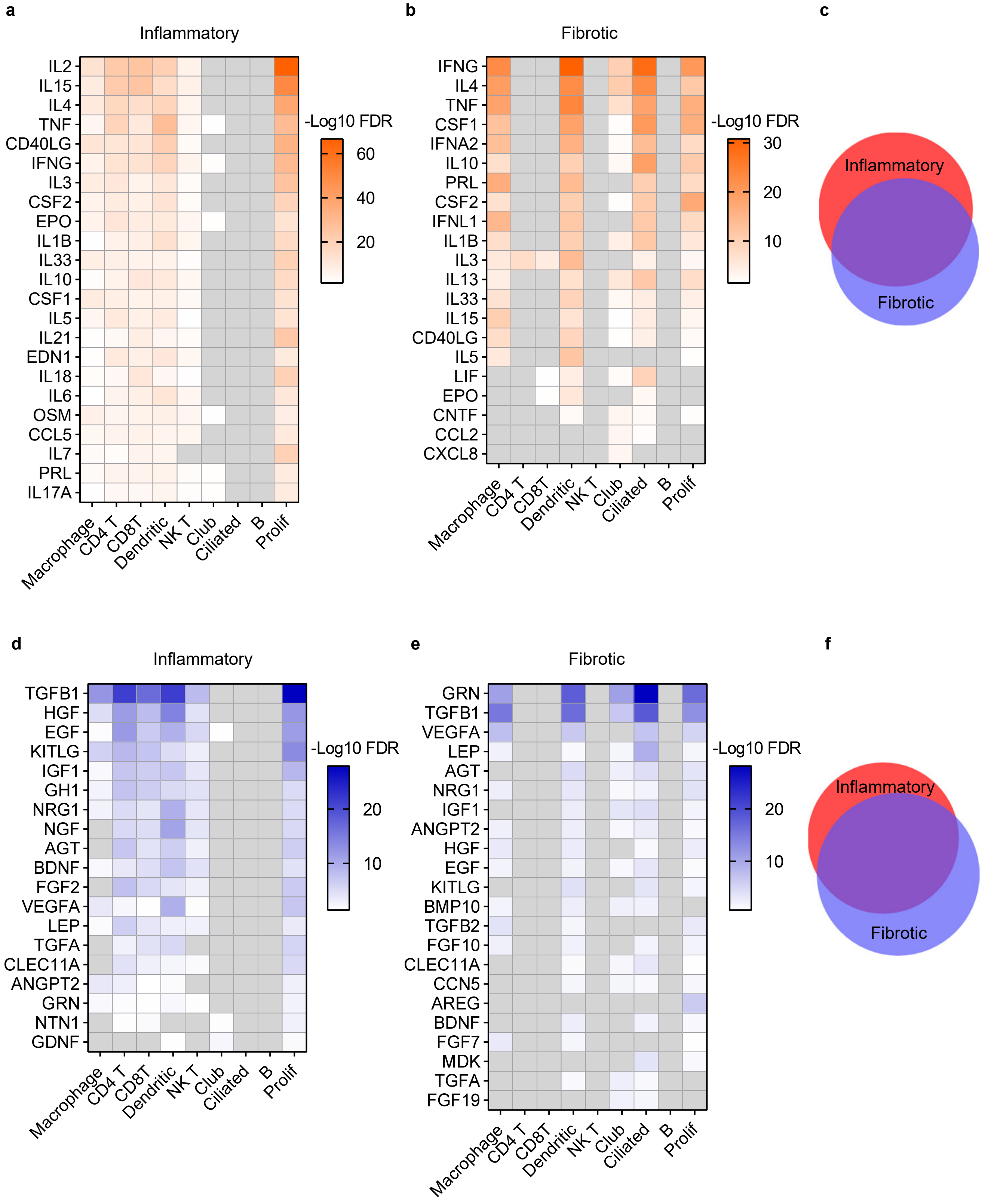

Supplementary Figure 4

**a**

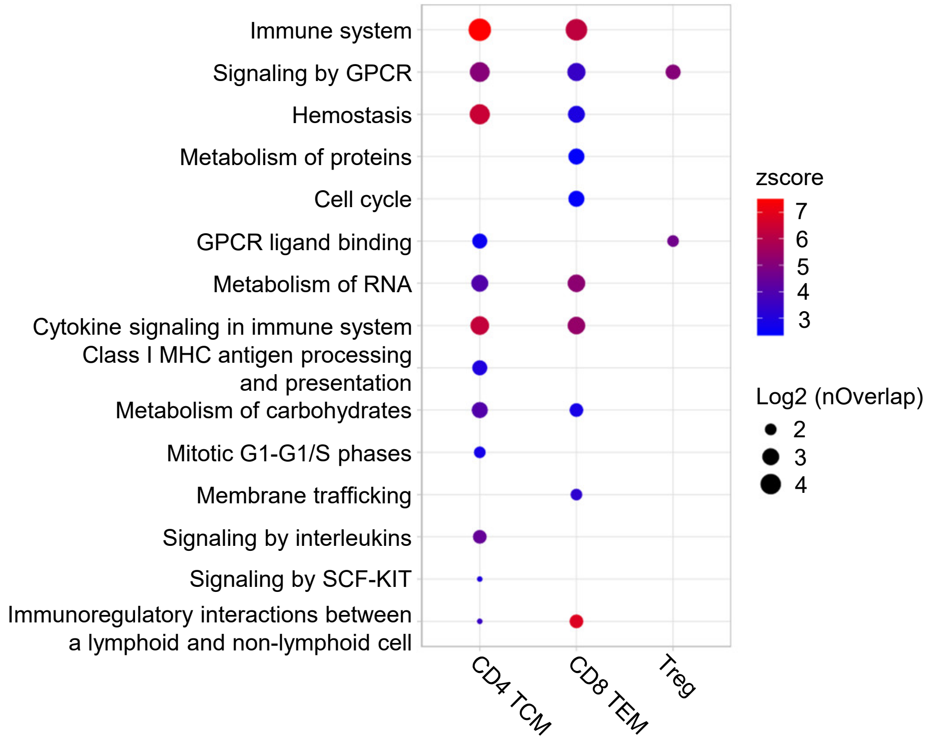

**b**

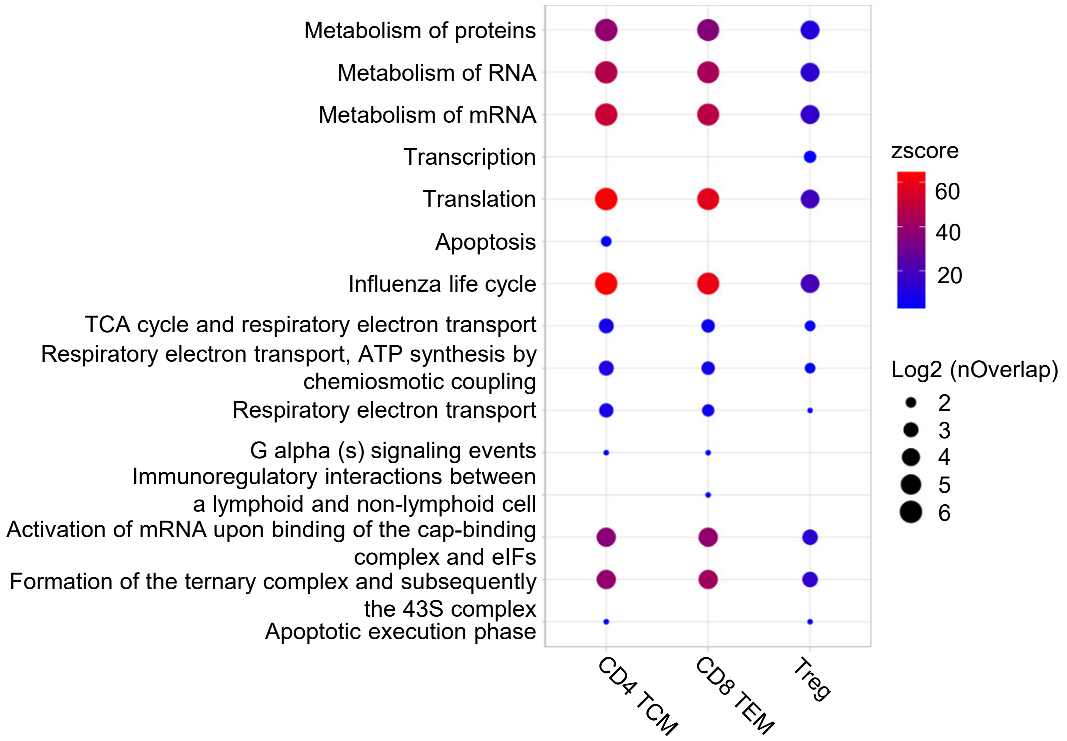

Supplementary Figure 5

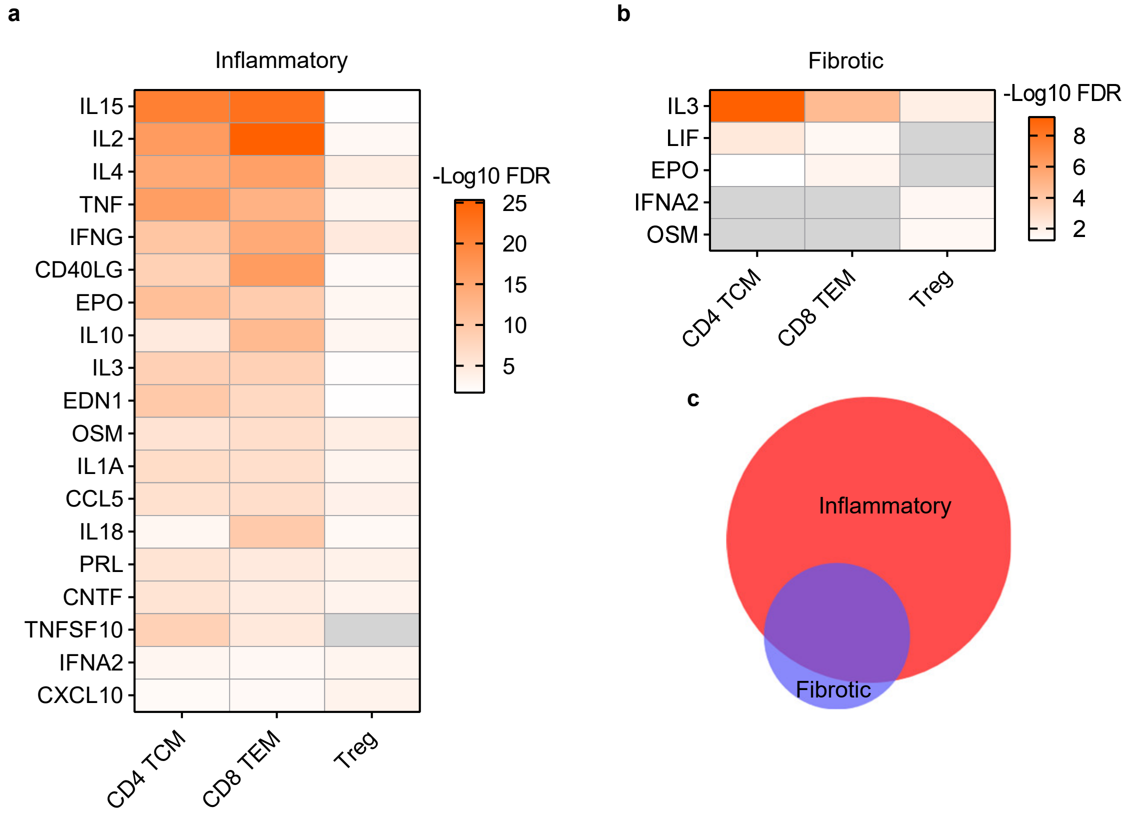

Supplementary Figure 6

a

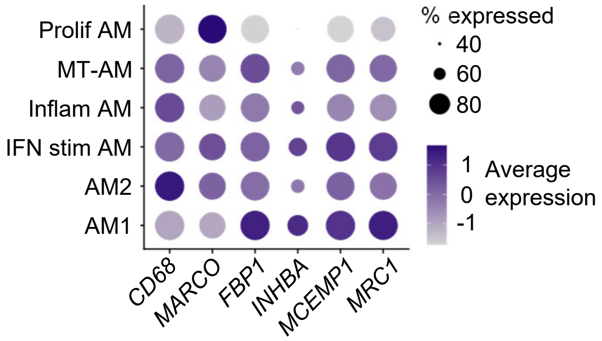

b

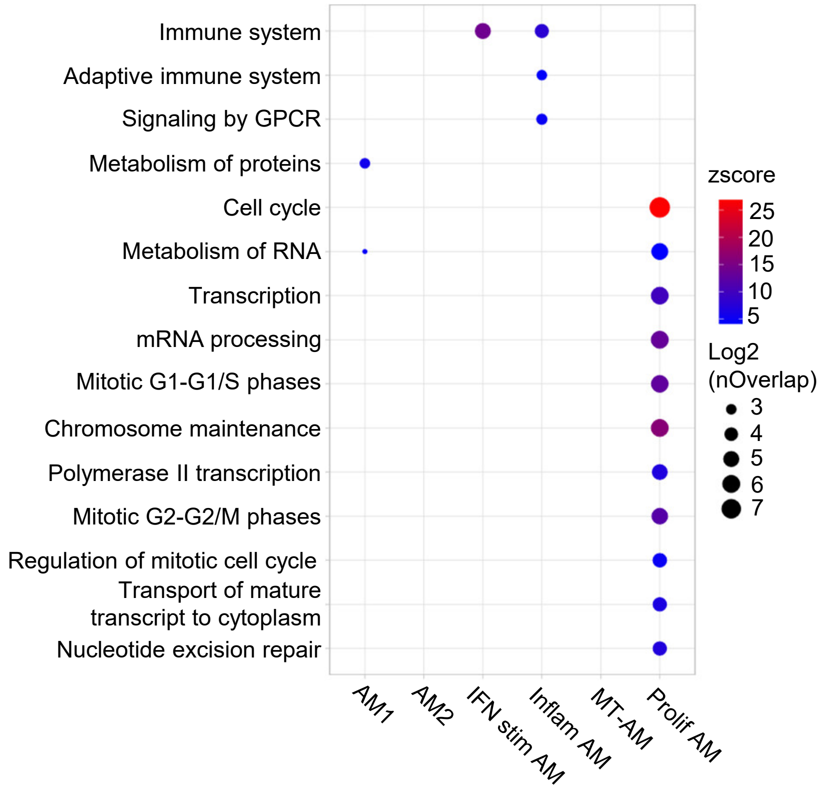

Supplementary Figure 7

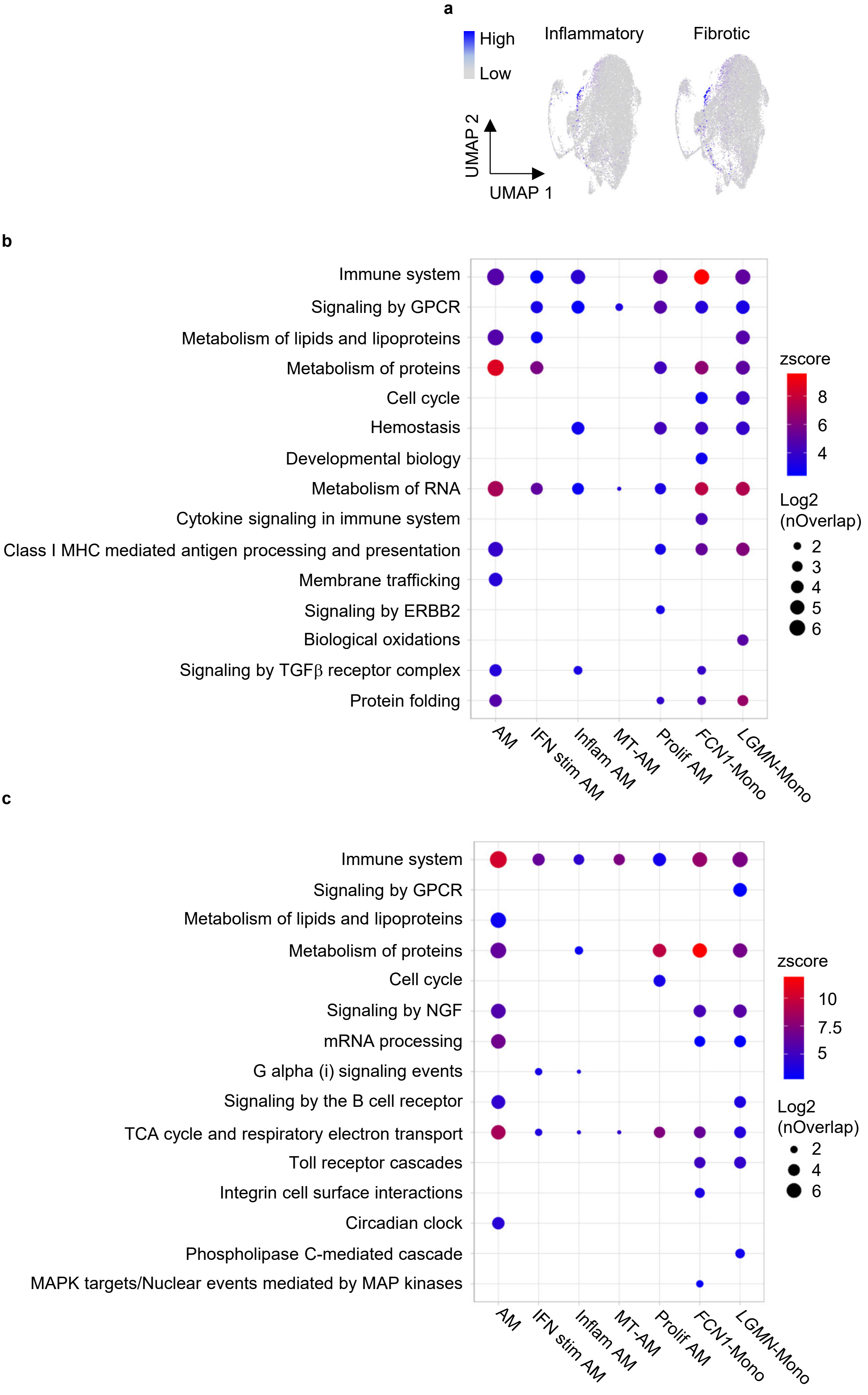

Supplementary Figure 8

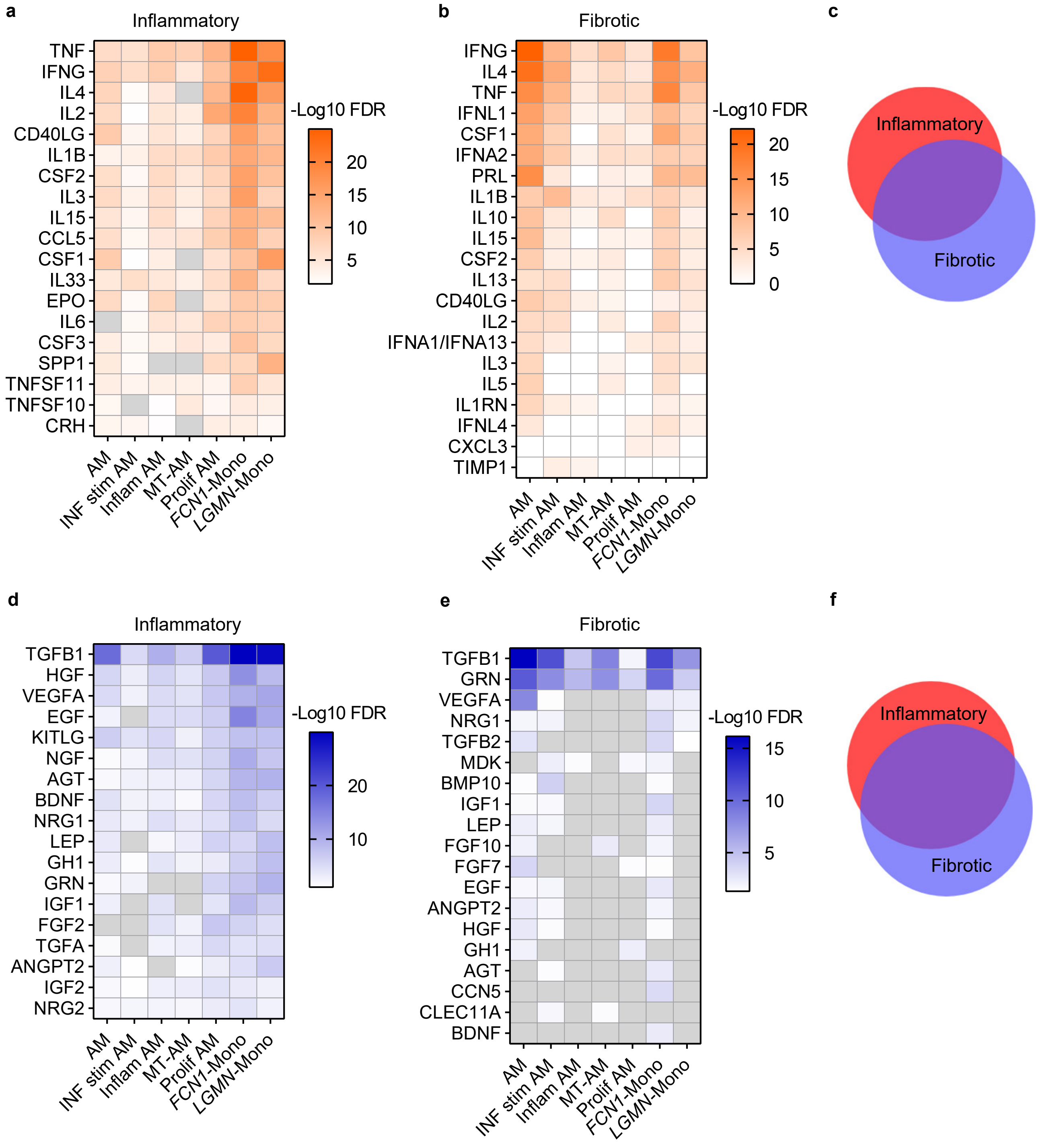
